# Prediction of etiology and prognosis based on hematoma location of spontaneous intracerebral hemorrhage

**DOI:** 10.1101/2024.05.22.24307743

**Authors:** Jingjing Liang, Weixiong Tan, Shijia Xie, Lijuan Zheng, Chuyan Li, Zhong Yi, Jianrui Li, Changsheng Zhou, Zhang Zhiqiang, Zhen Zhou, Ping Gong, Xingzhi Chen, Longjiang Zhang, Xiaoqing Chen, Qirui Zhang, Guangming Lu

## Abstract

**Background:** The characteristics of the hemorrhagic location of spontaneous intracerebral hemorrhage (sICH) is pivotal for both identifying its etiology and prognosis. While empirical conclusions have been obtained in clinical practice, a comprehensive and quantitative modeling approach has yet to be thoroughly explored.

**Methods:** We employed lesion-symptom mapping to extract the location features of sICH. We registered patients’ non-contrast computed tomography image and hematoma masks with standard human brain templates to identify specific affected brain regions. Then, we generated hemorrhage probabilistic maps of different etiologies and prognoses. By integrating radiomics and clinical features into multiple logistic regression models, we developed and validated optimal etiological and prognostic models across three centers, comprising 1162 sICH patients.

**Results:** Hematomas of different etiology have unique spatial distributions. Location features robustly categorized the etiology sICH (mean area under the curve (AUC) = 0.825) across different datasets), yielding clear add on value in models (fusion model mean AUC = 0.915) compared to clinical features (mean AUC = 0.828). In the prognostic analysis, patients with more extensive hematoma had a worse prognosis, the location (mean AUC = 0.762) and radiomic features (mean AUC = 0.837) also providing reliable add on value effects (fusion model mean AUC = 0.873) compared to clinical features alone (mean AUC = 0.771).

**Interpretation:** Our results show that location features were more intrinsically robust, generalizable relative, strong interpretability to the complex modeling of radiomics, our approach demonstrated a novel interpretable, streamlined, comprehensive etiologic classification and prognostic prediction framework for sICH.

## Introduction

Spontaneous intracerebral hemorrhage (sICH) is characterized by the extravasation of blood within the intracranial space due to the rupture of cerebral vessels, encompassing brain parenchymal hemorrhage, intraventricular hemorrhage, and subarachnoid hemorrhage. The heightened mortality rate and unfavorable prognosis associated with sICH poses a significant challenge for clinicians and patients. Non-contrast computed tomography (NCCT) imaging is the preferred method of diagnosis^1^. Within the context of sICH (nontraumatic), the identification of disparate etiologies gives rise to distinct and varied treatment indications. For instance, aneurysms may require interventions such as embolism or craniotomy, while hypertensive intracerebral hemorrhage is typically addressed through conservative or decompressive measures. Upon hospital admission the primary imperative for sICH patients is prompt determination of the underlying cause^2^. Survivors of sICH frequently encounter severe complications, enduring neurological impairments, even debilitating disabilities, if not treated promptly and correctly^3, 4^.

In their quest to identify the underlying etiology of sICH, clinicians routinely incorporate brain spatial location into their assessments of hemorrhagic impact. For instance, aneurysmal hemorrhage tends to manifest in the subarachnoid space^5^, hypertensive hemorrhage predominantly presents in the deep nucleus region^6^, and vascular malformations are commonly observed in cortex^7^. And in terms of prognostic prediction, hyperglycemia, hypertension, advanced age, and hematoma location characteristics, all have been empirically identified as crucial factors that influence outcomes^3, 8^. Accordingly, the localization of hematoma is of paramount importance for both etiologic and prognostic purposes.

Current analyses of sICH location often focus on specific cerebral regions, lacking a comprehensive investigation across the entire anatomical space of the brain. Lesion-symptom mapping, a population-based analysis method, provides standardized location features of lesions by calculating normalized lesion locations. In recent years, this method has been used in studies of ischemic stroke and glioma, yielding significant advances in the precision and full context of localization^9–12^. Utilizing this approach, probabilistic maps can be drawn, and the impact of clinical variables can be statistically quantified, enabling a quantitative analysis of location features based on the visualized distribution of hematoma effects throughout the entire brain^13, 14^.

Based on these insights, we posit that the location features of hematoma bear a close association to etiology and possess predictive value for determining prognosis. In this project, we employed lesion-symptom mapping to extract the location features of sICH and generated probability maps of etiology and prognosis for each and every brain region. We then combined these radiomic metrics with clinical features (e.g. hypertension, sex, and chronological age) to train and multiple models of etiology and prognosis, utilizing model comparison statistics to determine final, optimal models. Our study encompassed a comprehensive cohort of 1162 sICH patients across three medical centers, ensuring a robust, diverse, and generalizable dataset for our analyses. This study’s fundamental aim is to expand our understand how lesion location features affect the etiology and prognosis of sICH, and to provide a quantitative methodology for efficiently making these determinations.

## Materials and Methods

### Study Population and clinical Information

This retrospective study encompassed the collection of sICH patients from three distinct Yi Jishan Hospital (dataset 2, January 2018 to December 2020), and Nanjing First Hospital (dataset 3, January 2019 to December 2020). The confirmation of intracranial hemorrhage was achieved through NCCT. Patients were excluded if they presented with secondary hemorrhage resulting from head trauma, hemorrhagic transformation of ischemic infarction, brain tumors, or exhibited abnormalities in blood coagulation, liver, kidney function, or were subject to drug-induced cerebral hemorrhage. Supplementary Figure 1 illustrates the retrospective inclusion and exclusion flowchart in detail.

The following baseline clinical characteristics of patients were extracted from medical records: chronological age, sex, hypertension (defined as systolic blood pressure ≥140 mm Hg and/or diastolic blood pressure ≥90 mmHg, or self-reported use of antihypertensive medication within the past 2 weeks), diabetes mellitus (fasting plasma glucose concentration ≥7.0 mmol/L or exceeding 11.1 mmol/L 2 hours post-meal), hyperlipidemia (abnormal blood lipids were detected in serum), history of previous stroke, smoking, alcohol consumption, Glasgow Coma Scale (GCS) score at admission, and the known time interval from onset to baseline CT if witnessed or self-reported by the patient. All demographic and clinical information are shown in Table 1.

**Table 1.** Demographic and clinical characteristics in sICH patients.

| **Variables** | **Subjects (n = 1162)** |
| --- | --- |
| Age, years, mean ± | 57.65 ± 13.19 |
| GCS, median (IQR) | 12 (10-14) |
| Sex, n (%) |  |
| Male | 655 (56.4%) |
| Female | 507 (43.6%) |
| Hypertension, n (%) | 791 (68.1%) |
| Diabetes mellitus, n | 110 (9.5%) |
| Hyperlipidemia, n (%) | 27 (2.3%) |
| Smoking, n (%) | 220 (18.9%) |
| Drinking, n (%) | 219 (18.9%) |
| Onset-to-CT time, n |  |
| $\leq 24$h | 583 (50.2%) |
| 24h-3d | 428 (36.8%) |
| 3-14d | 128 (11.0%) |
| $>14$d | 23 (2.0%) |
| History of stroke, n | 93 (8.0%) |
| Etiology, n (%) |  |
| Hypertension | 563 (48.5%) |
| Aneurysm | 435 (37.4%) |
| Vascular malformation | 111 (10.0%) |
| Unknown | 53 (4.6%) |
| Prognosis, n (%) |  |
| mRS$\leq 2$ | 721 (62.1%) |
| mRS$>2$ | 441 (37.9%) |

GCS= Glasgow Coma Scale; mRS= the modified Rankin Scale

The etiologies of sICH were categorized into four classifications: hypertension, aneurysm rupture, vascular malformation, and clinically unclear diagnosis by discharge diagnosis. Modified Rankin Scale (mRS)^15^ scores were determined based on clinical examinations or telephone follow-ups at twelve months post sICH event. Consistent with prior literature, we defined good outcome as “mRS = 0-2”, poor outcome as “mRS = 3-6”^16, 17^

Ethics review and institutional review board approval in each center were both obtained. Informed consents from participating patients were exempted because of the retrospective nature of this study.

### Image Acquisitions and Preprocess

NCCT covering the entire brain was acquired with a slice thickness of less than 10.0 mm in all patients. All images underwent meticulous quality checks to eliminate those displaying significant motion artifacts. The hematoma areas were delineated from the NCCT images utilizing artificial intelligence brain hemorrhage auto-segmentation software (Dr. Wise TM software available at http://label.deepwise.com/). A physician’s review of the segmentation results revealed no obvious segmentation errors.

### Lesion-symptom mapping and location features

The NCCT scans and hematoma areas were subjected to spatial normalization and alignment with the standard Montreal Neurological Institute (MNI) template. The normalization process employed the CT normalization function offered by Standard Space Lesion Symptom Mapping (SPLSM, https://github.com/JLhos-fmri/SPLSM_version1.0) toolkit, which was based on the Clinical Toolbox of SPM12 (https://github.com/spm/spm12), utilizing a cost function masking method. Based on normalized hemorrhagic areas, the frequency occurrence of hematoma at each voxel was computed, creating a voxel-based probabilistic map for each patient (Figure 1A).

**Figure 1.**
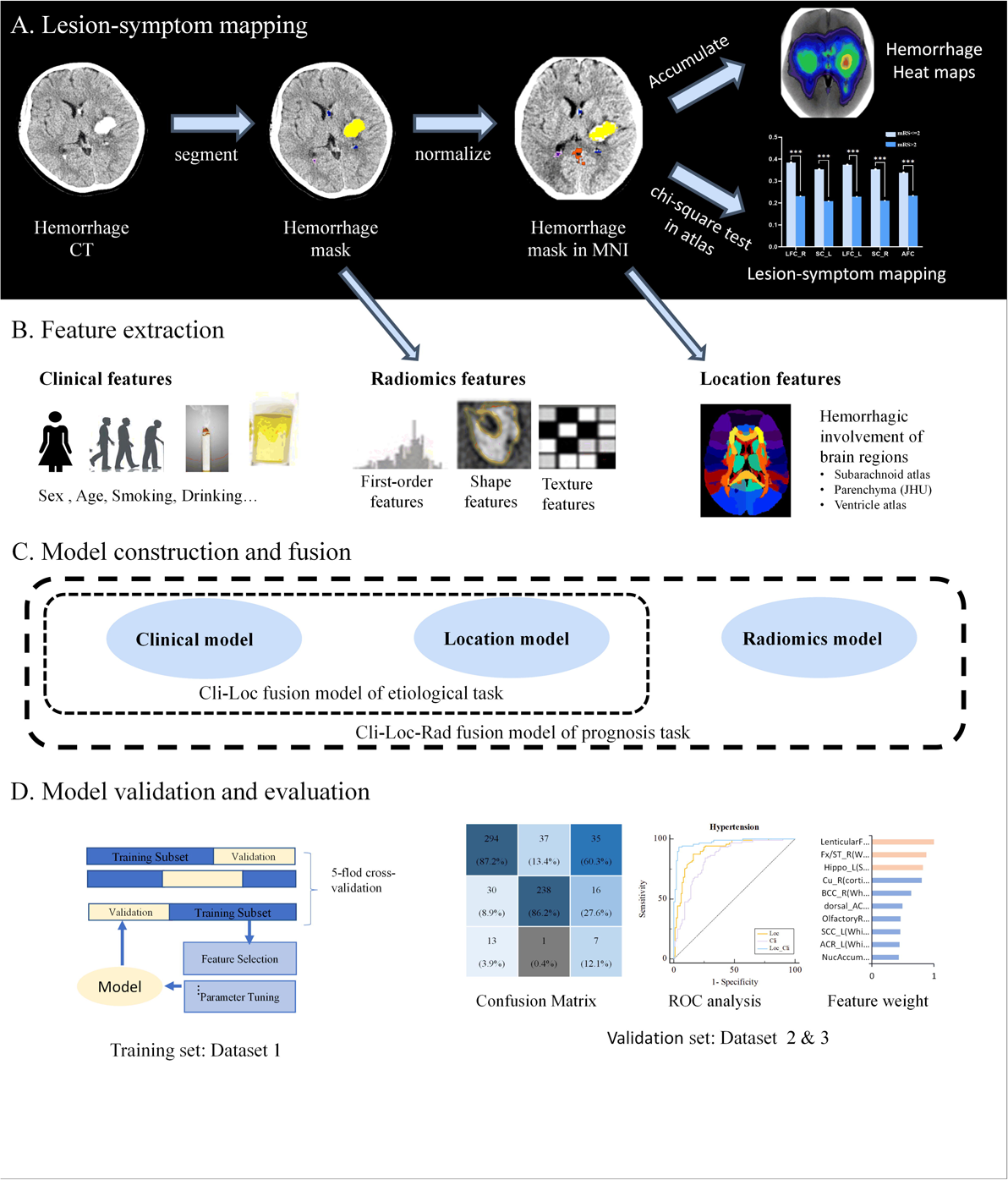
The workflow of constructing clinical-, location-, radiomics- and fusion models for prediction diagnosis of sICH. (**A**) Lesion-symptom mapping and location features. (B) Clinical, imaging and location feature extraction. (**C**) Construction and fusion of etiologic model and prognostic model. (D) Model validation and evaluation. ROC= Receiver operating characteristic.

This yielded a location feature set that quantified hemorrhage involvement of intracerebral regions in the parenchyma (cortical, white matter, and sub-cortical), subarachnoid space, and ventricles. A hemorrhagic event was considered present if more than 20% of the voxels in a given region showed a positive indication of a hemorrhage.

The brain parenchyma analyses utilized the Johns Hopkins University (JHU) template, comprised of 178 annotated brain regions, encompassing 68 white matter and 110 gray matter regions (Figure 1B). The subarachnoid space (10 regions) and ventricles (4 regions) were delineated using an in-house manually crafted template. List of intracranial regions see Supplementary Table 1.

In the etiological prediction analysis, 192 regions were sorted into 5 intracranial structures: ventricle, subcortical, white matter, cortical, and subarachnoid space. In each intracranial structure, a paired one-way ANOVA was used to calculate whether there was a significant difference in the rate of involvement in the region of the hemorrhage for the three etiologies. In the prognostic analysis, the rate of hemorrhage at 192 regions were compared by Chi-square test.

### Radiomics features

Radiomics features were extracted by the python package PyRadiomics (Version 3.0.1; https://pypi.org/project/pyradiomics). 1454 radiomic features were extracted for each hematoma region. It included first-order features, shape-based features, gray cooccurrence matrix, gray dependence matrix, gray running length matrix, gray size area matrix, and a neighborhood gray-tone difference matrix (Figure 1B). Given the possibility of multiple regions of hematoma of a portion of patients, it was essential to assign weights to the radiomic features of each region, creating a patient-level feature set. To achieve this, we used a deep learning multi-locus attention model algorithm to generate adaptive weights of focus-level features. The model underwent training and iteration within a training cohort using 5-fold cross-validation. The adjustment of model parameters occurred based on prognostic and error considerations (loss function). Subsequently, the radiomic features from multiple regions were amalgamated into patient-level radiological group features, as detailed in the Supplementary Method and Supplementary Figure 2. After adaptive weighting, 1454 radiomics features based on patient level were obtained.

### Model development for etiology and prognosis

The models developed in this study served dual purposes: classifying sICH etiology and predicting prognosis. In the actual clinical determination of etiology, doctors often judge by clinical characteristics and the location of hemorrhage. Therefore, we only included cases with known clinical and hemorrhage location features in our etiology classification models (n.b., by this criterion 53 patients with an unknown etiology at discharge were excluded). A multiclassification strategy was also used to predict whether the etiology is hypertension, ruptured aneurysm, or vascular malformation. Three models were constructed: clinical features only, location features only, and a fused location-clinical model (referred to as the loc-cli model).

In the prognostic task, clinical and hemorrhage location features, along with the above 1454 radiomic features, were utilized. Based upon these feature sets used to predict good (mRS = 0-2) or bad (mRS = 3-6) outcome^16, 17^. Four models were tested: clinical, location, radiomics, and a combined or fused clinical-location-radiomics model (referred to as the loc-cli-rad model) (see Figure 1C). Dataset 1 was utilized for model training, while datasets 2 and 3 were reserved for independent testing and validation of the models.

Logistic regression in scikit-learn (https://scikit-learn.org) toolkit was used to construct the model. In training set, Spearman correlations were used to eliminate feature redundancy. Correlation coefficients greater than 0.8 between two or more features are retained for only one of them. L1 feature selection methods were adopted for reducing dimensionality in our models. Fivefold cross-validation and a grid search with F1 score as the optimization goal were implemented on the training set, thereby tuning the model hyperparameters and predictive performances. The fusion model utilized “model fusion” in the training set, a technique that employed an average information strategy to combine the prediction scores generated after running each individual model during cross validation.

In modeling etiology, we plotted confusion matrices and receiver operating characteristic (ROC) curves to evaluate the models. To obtain ROC curves from these tri-categorical models, we reduced the multiclassification problem to three two-level classification problems, and averaged the corresponding true positive rate at the same false positive rate on the ROC curve and plotted the macro average ROC curve using the average of false positive rate and true positive rate.

In the prognosis prediction model, ROC curves were used for model evaluation. To compare the accuracy between models, we calculated the difference in the area under the curve (AUC) values between the fusion model and the other models. Subsequently, we verified the significance of these differences using the Delong test (Figure 1D).

## Results

This study involved a total of 1162 patients (655 males, 507 females), with a mean age of 57.65 ± 13.19 years. We categorized sICH etiology as hypertension, aneurysm, vascular malformation, or unknown, with respective counts of 563, 435, 111, and 53 patients. The clinical functional prognosis was stratified by outcome, with 721 and 441 patients for good and poor outcomes, respectively. Table 1 provides a comprehensive summary of sample demographic and clinical characteristics. Detailed demographics and clinical characteristics for each dataset are provided in Supplementary Results, Supplementary Tables 2 and 3.

### Association between hematoma location and etiology

The distribution of hematoma by etiology (hypertension, aneurysm, vascular malformation) is depicted in Figure 2A. Hypertensive hematoma manifested primarily in deep brain, particularly the basal ganglia area. Conversely, hematoma resulting from aneurysm rupture were concentrated primarily in the subarachnoid space, encompassing regions such as the lateral fissure cistern and the anterior longitudinal fissure cistern in subarachnoid area.

**Figure 2.**
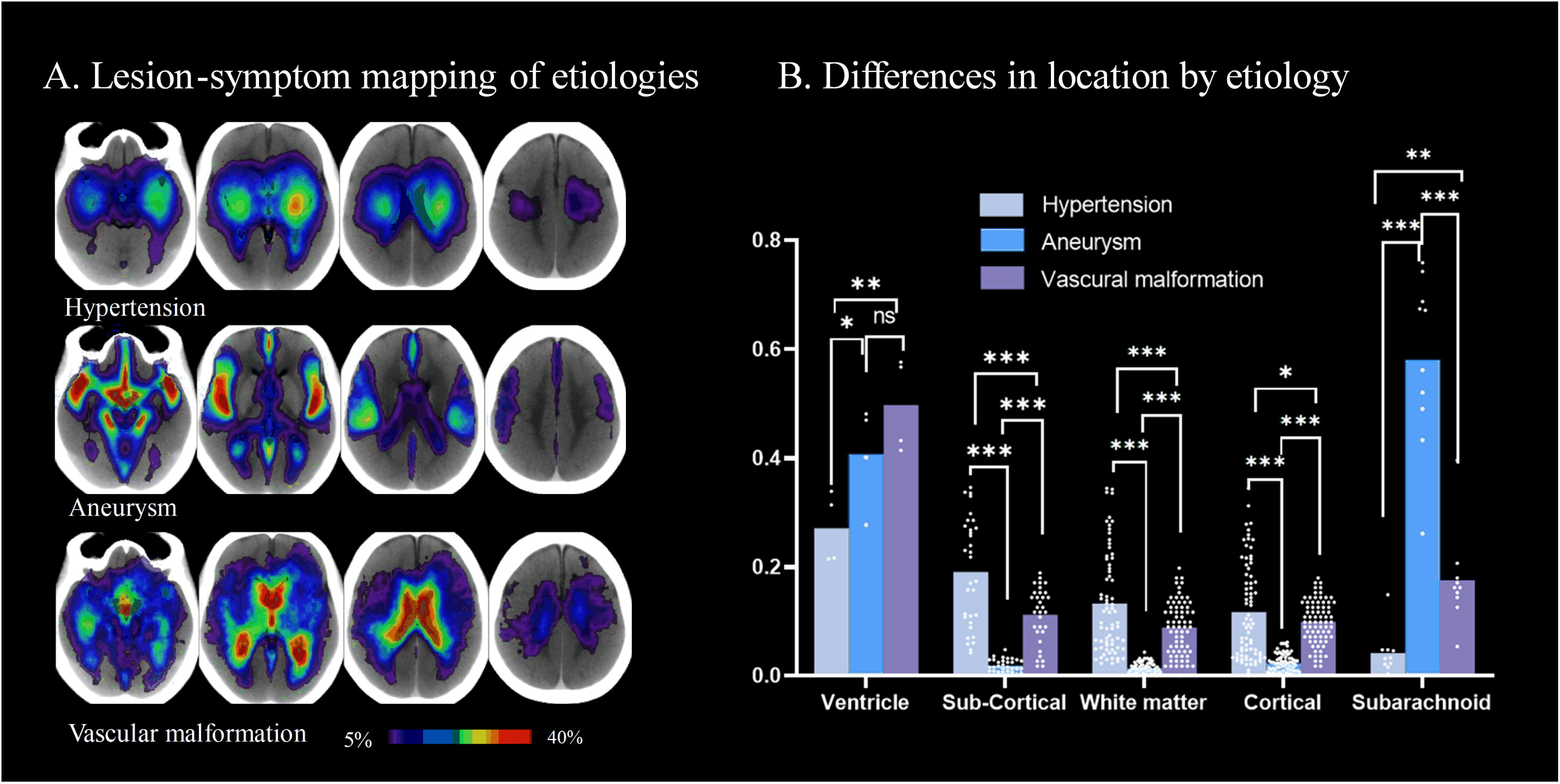
Association between hematoma location and etiology (n = 1109). (A) Probability maps depicting the distribution of hematoma location in patients. The location patterns associated with hypertension, aneurysm, and vascular malformation are illustrated, with the frequency of hemorrhage location graded according to color. The gradient scale ranged from 5% to 40%. (B) Paired sample ANOVA was used to compare the involvement rate of each region in each intracranial structure. *: p < 0.05, **: p < 0.01, ***: p < 0.001, ns: p > 0.05.

Notably, vascular malformation-related hematoma demonstrated a more even and widespread distribution, with a higher likelihood in the ventricles. The proportion of individuals affected in each intracranial region based on intracranial structure (ventricle, subcortical, white matter, cortical, and subarachnoid space) is summarized in Figure 2B. Significant differences were observed in the distribution of cerebral hemorrhage for the different etiologies. A noteworthy result is that vascular malformations and ruptured aneurysms appeared likely to involve the ventricles. Hypertension and vascular malformations tended to involve the brain parenchyma, but hypertension tended to be sub-cortical, whereas vascular malformations tended to be cortical. In addition to ruptured aneurysms, which are most likely to involve the subarachnoid space, vascular malformations also involved the subarachnoid space. The pattern of hematoma distribution was consistent across datasets 1, 2, and 3 (Supplementary Figure 3, 4, 5).

### Association between hematoma location and prognosis

Probability plots show that, in general, patients with a poorer prognosis have a wider distribution of hematoma in subarachnoid space, parenchyma, and ventricles (Figure 3A). When the hematoma was located in the radiation crown, marginal gyrus, cerebellum, bilateral lateral ventricle and fourth ventricle, the prognosis was generally poor. In contrast, patients with hematoma in the subarachnoid area had a relative good prognosis (Figure 3B). Detailed results for the separate datasets (1, 2, and 3) are provided in Supplementary Figure 6, 7, 8.

**Figure 3.**
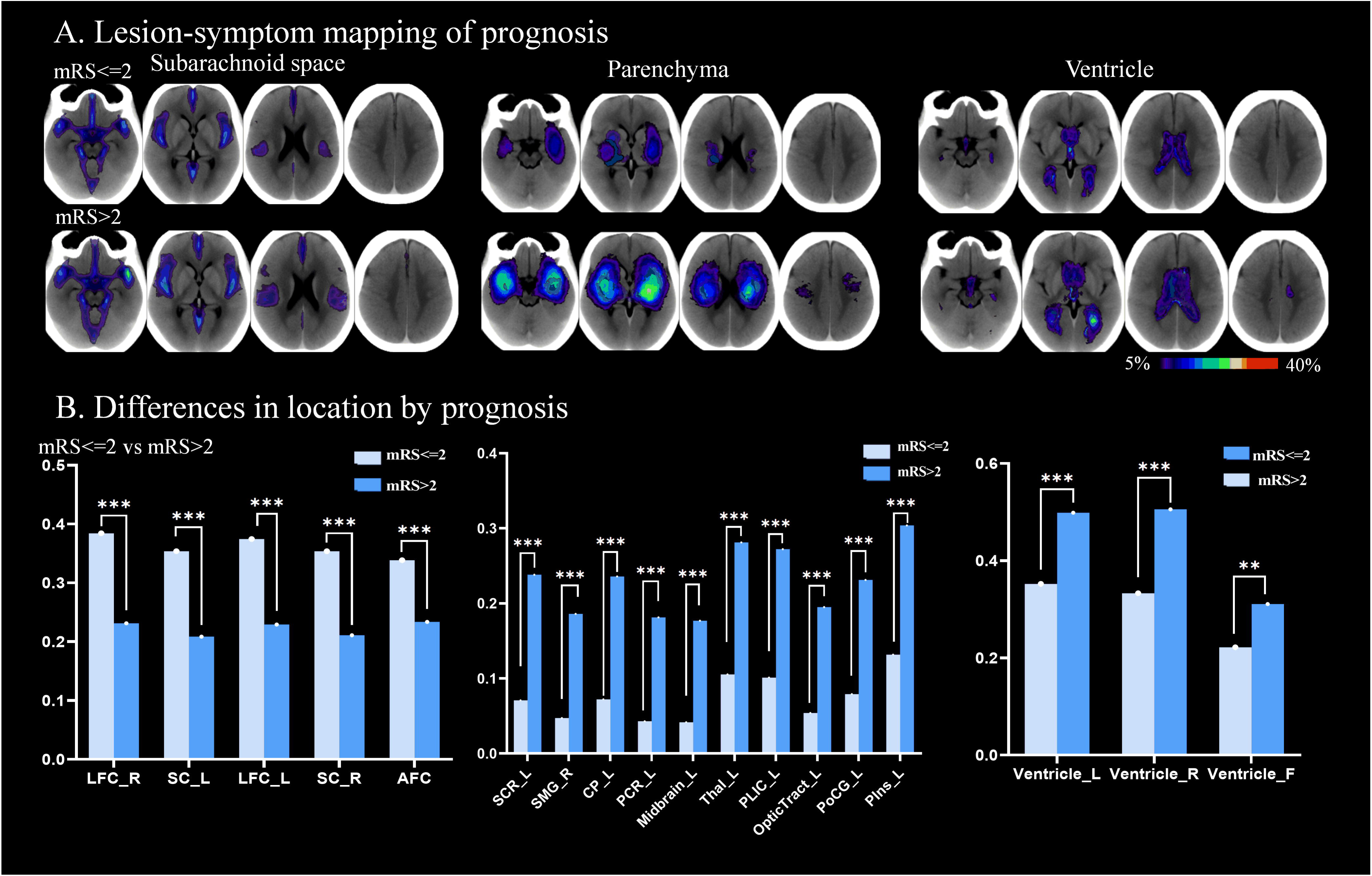
Association between hematoma location and prognosis (n = 1162). (**A**) Probability maps depicting the distribution of hematoma location in different prognosis. The location patterns associated with subarachnoid space, parenchyma and ventricle are illustrated, with the frequency of hematoma location graded according to color. The gradient scale ranged from 5% to 40%. (**B**) Frequency plots of the top five cisterns, ten parenchyma and three ventricles showing a group difference are ranked according to chi-square values. *: p < 0.05, **: p < 0.01, ***: p < 0.001, ns: p > 0.05. Abbreviations of intracranial region see Supplementary Table 1.

### Etiology classification

In the etiological classification task, the clinical model demonstrated an average AUC of 0.828 (range: 0.806 ∼ 0.848 cross different dataset), while the location model achieved an AUC of 0.825 (range: 0.807 ∼ 0.842). Notably, the fusion model exhibited a higher AUC of 0.915 (range: 0.909∼0.923). See Supplementary Table 4 for details of model performances in each dataset. The confusion matrix in Figure 4A was employed to assess the mean accuracy of each model cross datasets. The location models achieved overall accuracies exceeding 86% for hypertension and aneurysm, with suboptimal performance (12.6%) for the prediction of vascular malformation. The fusion model, however, improved the diagnostic ability of all three etiological types (hypertension = 94.7%, aneurysm = 91.5%), especially for vascular malformations (42.3%). Detailed results of each dataset are provided in Supplementary Figure 9. Delong tests confirmed that the fusion model consistently outperformed the clinical models, showing significantly higher AUC in all datasets (all p<0.001) (Supplementary Table 10), indicating that location features provided significant predictive value (Supplementary Table 5).

**Figure 4.**
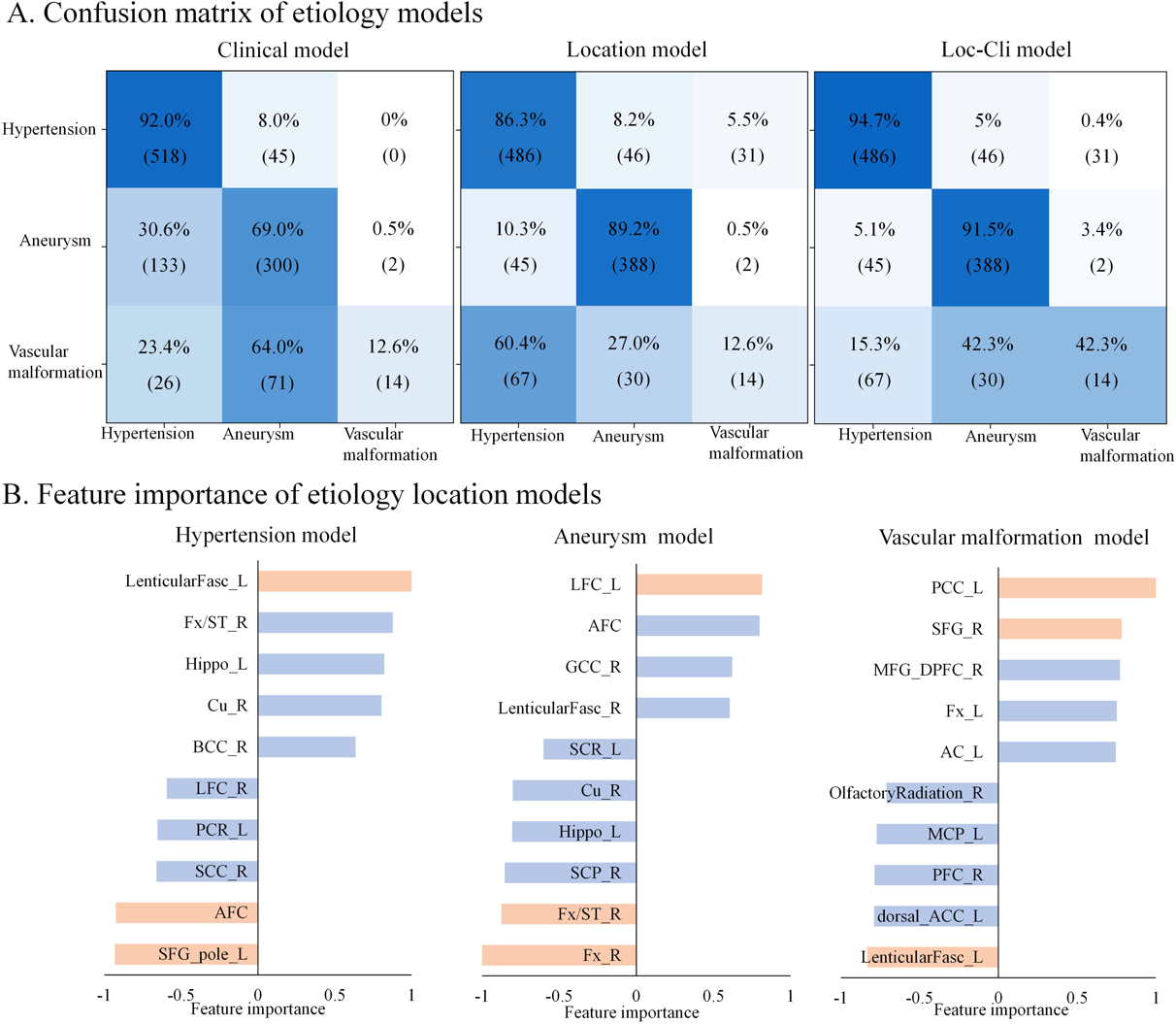
Etiology classification. (A) The confusion matrix demonstrated the classification capability of the location model for predicting etiology in all datasets. Columns represent predicted classes, with rows representing ground truth. (B) The top ten location features associated with hypertension, aneurysm, and vascular malformation prediction models are shown. The horizontal axis represents the relative feature weights, with red coding the three features making the strongest contributions to the model. Positive values indicate a positive contribution, negative values a negative contribution. Abbreviations of intracranial region see Supplementary Table 1.

In the clinical model, hypertension, male sex, and history of stroke displayed the strongest positive contribution to predicting hypertensive sICH, whereas these features showed the strongest negative contribution in association with aneurysms, indicating that the presence of these features made it less likely that an aneurysm was present. In vascular malformation, male sex had a positive contribution, whereas hypertension and age had a negative contribution (Supplementary Figure 11). In the location models, hypertension was strongly associated with deep regions such as the lentiform nucleus, striae terminalis, and hippocampus. In contrast, hemorrhage of the superior frontal gyrus and anterior longitudinal fissure contributed negatively to the model, reducing the likelihood that hypertensive sICH was present. Aneurysms ruptured mainly in the subarachnoid space, especially in the lateral fissure cistern and anterior longitudinal fissure cistern. Hemorrhage in the fornix and cerebellum, however, contributed negatively to the model, indicating that aneurysmal cerebral hemorrhage were less likely to reside in these areas. Vascular malformations were primarily distributed in the posterior cingulate gyrus and superior frontal gyrus, whereas they were rare in the lenticular fasciclus and dorsal anterior cingulate regions (Figure 4B).

### Prognosis prediction

In the prognostic prediction task, the clinical model demonstrated an average AUC of 0.771(range: 0.762 ∼ 0.799), while the radiomics model achieved an average AUC of 0.837 (range: 0.831 ∼ 0.840). In contrast, the location model exhibited an average AUC of 0.760 (range: 0.785 ∼ 0.725). The fusion model outperformed all other models with an average AUC of 0.873 (range: 0.895 ∼ 0.850). ROC of each dataset is shown in Figure 5 Detailed model performances are shown in Supplementary Table 4. Delong tests confirmed that the fusion model consistently outperformed the clinical models, displaying significantly higher AUC in all datasets (all p < 0.01), indicating that location and radiomics features consistently provided predictive value above and beyond standard clinical predictors (see Supplementary Table 5).

**Figure 5.**
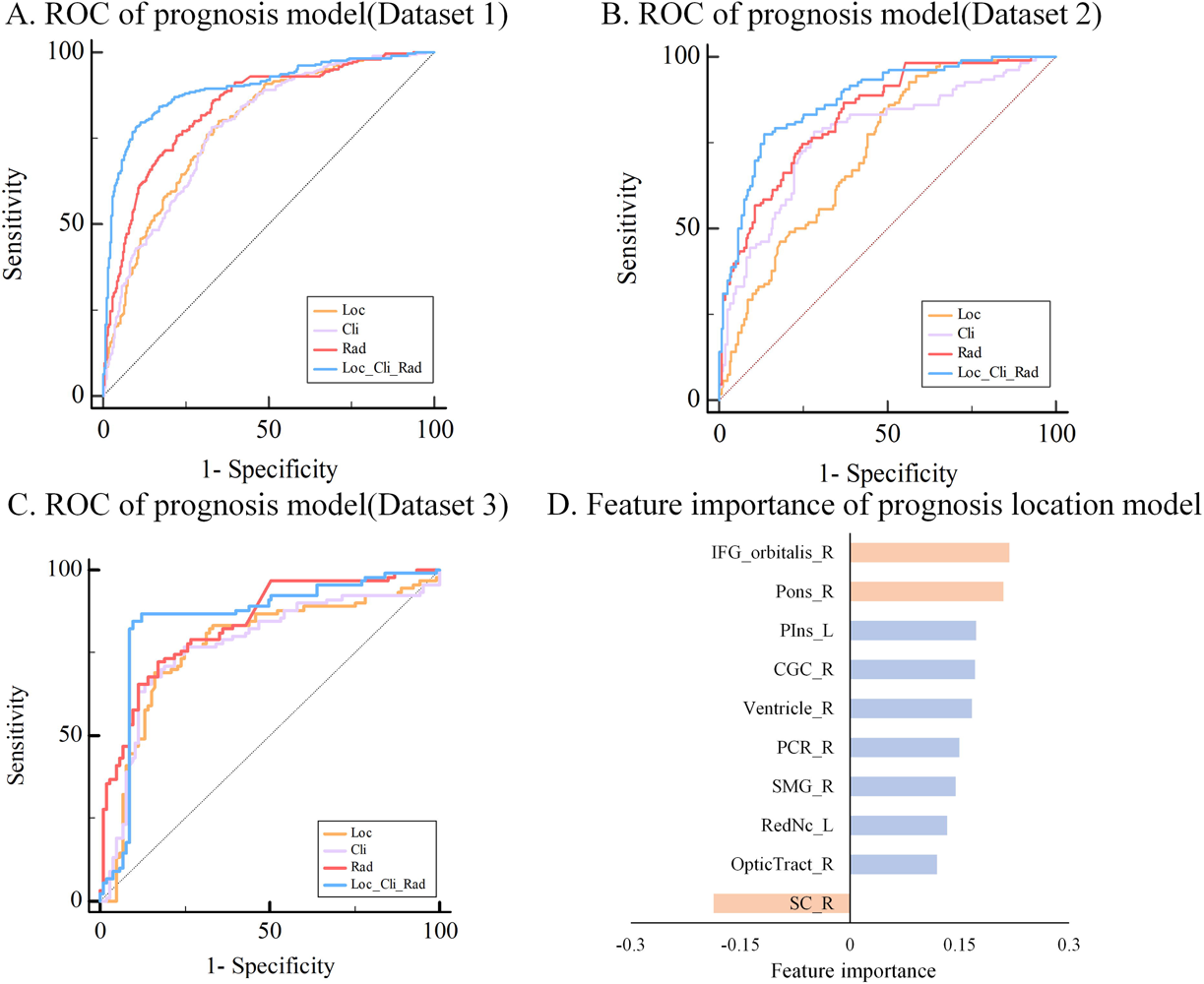
Prognosis prediction. (A-C) Receiver Operating Characteristic (ROC) curves of sICH predicted by the clinical, location, radiomicsand multidimensional fusion models of dataset1, 2 and 3. (D) displays the top ten location features of in the location model of prognosis prediction task. The horizontal axis represents the model weight value, red codes the three features making the strongest contribution. Abbreviations of intracranial region see Supplementary Table 1.

The most important features in the clinical model were lower GCS, age and presence of hypertension. In contrast, in the radiomics model, higher shape mesh volume original, lower GLDM, and lower NGTDM were the most important (see Supplemental Figure 12 for a complete list of feature weights). In the location model (see Figure 5D), the most important features making a positive contribution to poor prognosis were right inferior frontal gyrus orbitalis, right pontine, left posterior insula, and right cingulate gyrus (n.b., the suprasellar cisterna negatively contributed to poor prognosis).

## Discussion

This study provides a novel, interpretable, streamlined, comprehensive framework for etiologic classification and prognostic prediction of sICH patients after NCCT examination. In particular, we found that the hematoma location features in our models accurately described the relationship between spatial hematoma distribution and etiology or prognosis, with these relationships consistent with a priori clinical knowledge. This consistency suggested that imaging biomarkers constructed using hematoma location features are highly interpretable. As our imaging markers were not affected by different centers or different NCCT scanners, they appeared robust, exhibiting a high degree of generalizability. Lastly, our data showed our imaging biomarkers provided predictive above and beyond standard clinical characteristics. Accordingly, our data not only enhances our understanding of the condition, but showed the potential to improve clinical decision-making that would be broadly applicable.

Clinical features have always been important information for clinicians to judge the cause of intracerebral hemorrhage and clinical treatment^18^. In our clinical model, as expected, hypertension was crucial for hypertensive sICH, as hemorrhage often occurs in the setting of poor blood pressure control^19^. For aneurysm patients, female showed higher risk than male, it is consistent with previous studies, noting that additional hemorrhage from a ruptured aneurysm is more common in females^20, 21^. Our results also showed age of vascular malformations patients is younger than hypertension and aneurysm patients. This is due to the fact that brain hemorrhages caused by congenital dysplasia usually precede those caused by structural degeneration of vessels^22^.

On the basis of our clinical model, we utilized lesion-symptom mapping for quantitative and precise specification of hematoma location, confirming that location features have a significant additive effect relative to standard clinical information for the prediction and classification of etiology. Our data showed that hypertensive hemorrhage manifested primarily in deep brain regions, specifically the lentiform nucleus, striae terminalis, and hippocampus. This is consistent with the previous clinical knowledge, which contends that hypertensive hemorrhage is primarily located in the basal ganglia region^23^. Due to hemodynamic sensitivity, the cerebral arterial circle or deep penetrating arteries may exhibit heightened responsiveness to systemic blood pressure changes compared to cortical arteries^3, 24^, thereby increasing the likelihood of bleeding^25, 26^. Subarachnoid hemorrhage often results from rupture with sICH occurring in approximately 20-30% of aneurysm patients^27^. The cistern in which the hemorrhage is likely to be located is an important clue to finding the responsible aneurysm ^28, 29^ In this light, it is important to note that our etiologic classification model found evidence that involvement of the lateral fissure cistern and the anterior longitudinal fissure cistern were key in predicting aneurysm rupture. Our evidence also suggesting that the corpus callosum and lenticular fasciculus are also important in terms of location for regions outside the subarachnoid space. Vascular malformations are congenital vascular abnormalities^30^. Hemorrhages associated with vascular malformations can be distributed across the white matter, cortex, subcortex, subarachnoid space, and other regions, indicating that a broad range of potential bleeding sites. Our data demonstrate the unique exist in the posterior cingulate gyrus and superior frontal gyrus.

Numerous predictors associated with an elevated risk of adverse prognosis following sICH have been identified, encompassing clinical factors such as age, elevated blood pressure and blood glucose levels, as well as hematoma location, ^31, 32^. Our clinical model broadly align with these established associations. Compared with the young, older patients have vascular distortion, accompanied with atherosclerosis^33^, and this can co-exist with other diseases, all factors that can indirectly increase disability and mortality. Previous studies have found that uncontrolled blood pressure will increase the risk of stroke recurrence, and controlling the stability of blood pressure is crucial to preventing such events. Our data showed that GCS score is the strongest prognostic predictor ^34^ of sICH, with alow GCS score indicating severe disturbance of consciousness and a poor prognosis^35^.

When including location and radiomic features in our prognostic models, we found they provided predictive value significantly above what was provided by these more standard clinical features. In our prognostic models, brain regions located in areas of the cortex contributed more to the prognostic model, whereas subcortical regions contributed less to the prognostic prediction, which is consistent with previous studies that have concluded that lobar hemorrhages have a poor prognosis relative to nonlobar hemorrhages^32, 36^. The regions most associated with poor outcomes predominantly involve the inferior frontal gyrus, pontine, and insula. Damage to the inferior frontal gyrus has been shown to lead to language deficits^37^, may further hinder the recovery of physical function^38^. Pontine hemorrhage is particularly grave in terms of prognosis. After pontine hemorrhage, various oculomotor abnormalities (such as pupil apex, medial longitudinal tract syndrome, ocular deviation and nystagmus) may be observed^39^. The insula is a key node within the supratentorial swallowing network. Previous studies have shown a strong association between lesions of the right insular cortex and a severe symptom of dysphagia^40, 41^. At the same time, injuries involving the insula have also been found to increase cardiogenic mortality^42^.

Among the radiomic features, the shape mesh volume of the hemorrhage is positively correlated with prognosis, and this metric allows for a more precise characterization of the volume of irregular hemorrhage^43^. This is consistent with the common knowledge that the larger the hematoma, the worse the prognosis^44, 45^, and is also in line with our location-based analysis. The NGTDM features represent the contrast between voxels and neighboring voxels^46^, is used to quantify the intensity difference between neighboring voxels. The contrast which negatively contributes to prognosis in our data. This suggests that if the density is uneven due to the large volume of the hematoma, slow absorption or secondary bleeding, the contrast will be relatively low, which indicating a poor prognosis^47^.

At the level of feature construction, this study incorporates both and location features and radiomics features. Since location features are determined to describe the entire hemorrhagic region, they are not affected by NCCT scanning machines and parameters and are naturally compatible with the analysis of multiple hemorrhagic foci. In contrast, radiomics affected by the NCCT scanning machine and parameters^48^ and are difficult to deal with the combined analysis of multiple hemorrhagic regions. To address this issue, our approach incorporates innovative techniques for radiomics feature extraction. Through multiple instance learning, we assign weights to the cross-focus of each patient, ensuring the preservation of both global information pertaining to the patient’s lesions and the local information associated with each hemorrhagic location^49^. Even so, we believe that location features are intrinsically robust and generalizable relative to the complex modeling of radiomics, especially the high degree of interpretability that coincides with a priori knowledge.

Our study has several limitations. First, the study is a retrospective study, and the possibility of sample bias cannot be ruled out, e.g., the etiologic and prognostic distributions of the included samples may differ from the real situation. Second, we categorize the etiology of hemorrhage in general terms, which may reduce the applicability of our results in some cases affects the integrity of the etiological classification results. For example, sICH caused by amyloidosis is difficult to diagnose clinically, and there was no clear etiologic classification in our study.

In conclusion, our results show that the location features of hematoma are stable and reliable indicators in sICH etiology and prognosis prediction, and the combination of the radiomics features can effectively improve the prediction efficiency and overcome the defects of the instability of the radiomics features. Provide an interpretable, comprehensive etiologic classification and prognostic prediction framework for sICH in a streamlined manner.

## Supporting information

Supplemental files, and will be used for the link to the file on the preprint site.

## Data Availability

The data that support the findings of this study are available from the corresponding author on reasonable request

## Acknowledgements

We thank Prof. Joseph I. Tracy from Thomas Jefferson University for language editing and helpful discussions. This study received funding from National Natural Scientific Foundation of China (Grant Nos. 82271983 to X.C., 82127806 to G.L., 81790653 to G.L., and 81701680 to Q.Z.), National Key R&D Program of China (Grant Nos. 2020AAA0109505 to G.L., 2018YFA0701703 to Zhiqiang Zhang).

## Author Contributions

G.L., Q.Z., and X.C. contributed to the conception and design of the study; Q.Z. J.L., W.T. contributed to acquisition and analysis of data, drafting the text or preparing the figures; S.X., L.Z., C.L., Y.Z., J.L., C.Z., Zhiqiang Zhang., Zhen Zhou, P.G., X.C., L.Z. contributed to the acquisition and analysis of data

