## Supplemental files, and will be used for the link to the file on the preprint site. for "Prediction of etiology and prognosis based on hematoma location of spontaneous intracerebral hemorrhage": Supplementary Materials and Methods.docx

**Supplementary Methods**

**Study Population** **enrollment**

Inclusion and exclusion interflow are shown in Supplementary Figure 1.

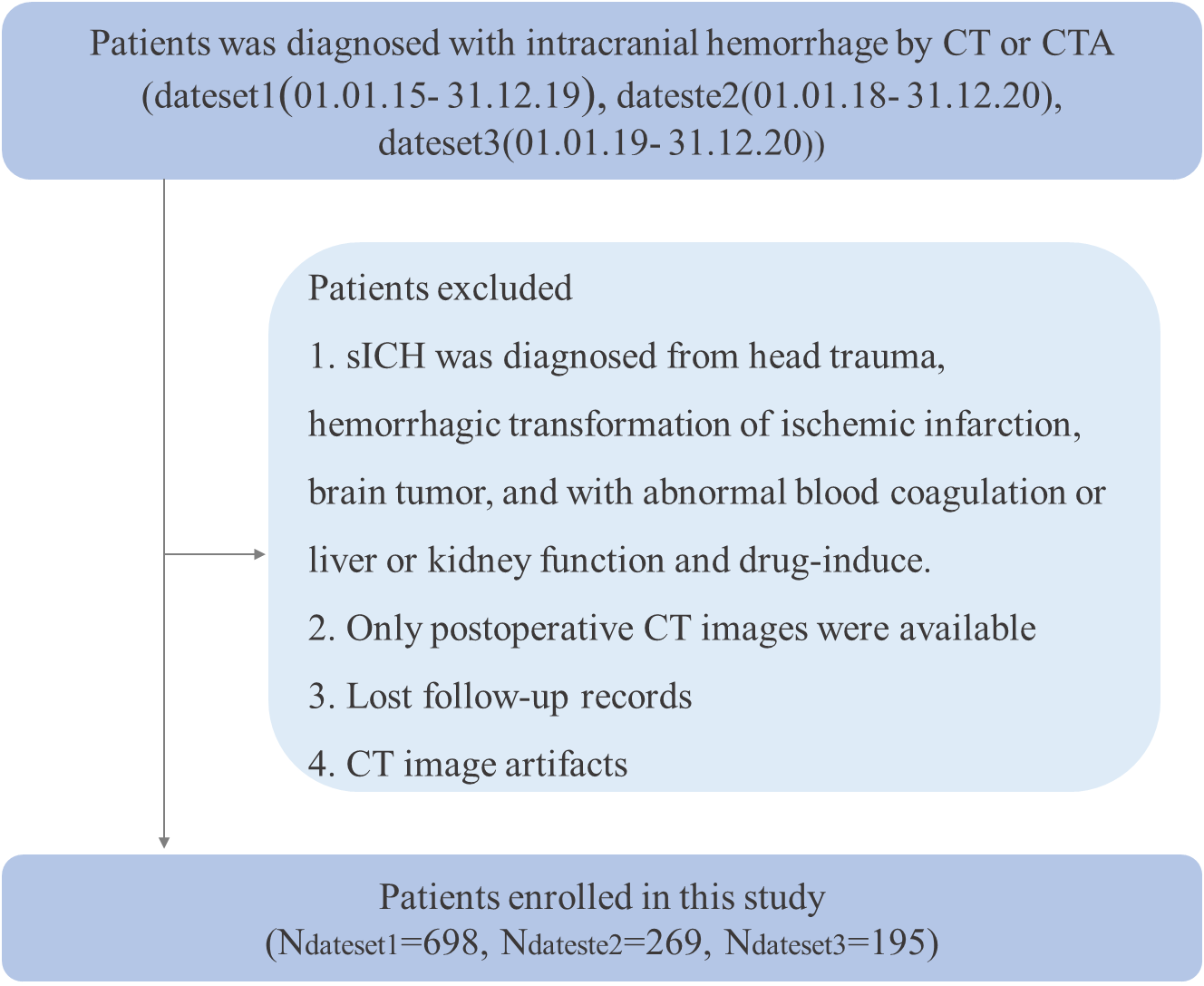
 **Supplementary Figure 1.** Flowchart shows the patient enrollment process.

dateset1 = Jinling Hospital, dateset2= Yi Jishan Hospital, dateset3 = Nanjing First Hospital

sICH= Spontaneous cerebral hemorrhage; CTA= Computer tomography angiography.

**Supplementary Table1.** **Intracerebral regions in the parenchyma, subarachnoid space, and ventricles.**

| **JHU template** | | **in-house template** | |
| --- | --- | --- | --- |
| **Abbreviation** | **Full title** | **Abbreviation** | **Full title** |
| Ins | insular | AFC | anterior longitudinal fissure cistern |
| Amyg | amygdala | PFC | posterior fissure cistern |
| Hippo | hippocampus | LFC | lateral fissure cistern |
| Caud | caudate nucleus | SC | suprasellar cistern |
| Put | putamen | AC | ambient cistern |
| GP | globus pallidus | PC | pontine cistern |
| Thal | thalamus | QC | quadrigeminal cistern |
| Hypothalamus | hypothalamust | Ventricle_L | left lateral ventricle |
| Mynert | nucleus innominata of mynert l | Ventricle_R | Right lateral ventricle |
| NucAccumbens | nucleus accumbens | Ventricle_T | third ventricle |
| RedNc | red nucleus | Ventricle_F | fourth ventricle |
| Snigra | substancia nigra |  |  |
| cerebellum | cerebellum |  |  |
| CP | cerebral peduncle |  |  |
| Midbrain | midbrain |  |  |
| PIns | posterior insula |  |  |
| CST | corticospinal tract |  |  |
| SCP | superior cerebellar peduncle |  |  |
| MCP | middle cerebellar peduncle |  |  |
| PCT | pontine crossing tract |  |  |
| ICP | inferior cerebellar peduncle |  |  |
| ML | medial lemniscus |  |  |
| Pons | pons |  |  |
| Medulla | medulla |  |  |
| ACR | anterior corona radiata |  |  |
| SCR | superior corona radiata |  |  |
| PCR | posterior corona radiata |  |  |
| GCC | genu of corpus callosum |  |  |
| BCC | body of corpus callosum |  |  |
| SCC | splenium of corpus callosum |  |  |
| TAP | tapatum |  |  |
| ALIC | anterior limb of internal capsule |  |  |
| PLIC | posterior limb of internal capsule |  |  |
| RLIC | retrolenticular part of internal capsule |  |  |
| EC | external capsule |  |  |
| CGC | cingulum (cingulate gyrus) |  |  |
| CGH | cingulum (hippocampus) |  |  |
| Fx/ST | fornix (cres) / Stria terminalis |  |  |
| Fx | Fornix |  |  |
| IFO | Inferior fronto-occipital fasciculus |  |  |
| PTR | Posterior thalamic radiation |  |  |
| SS | Sagittal stratum |  |  |
| SFO | Superior fronto-occipital fasciculus |  |  |
| SLF | Superior longitudinal fasciculus |  |  |
| UNC | Uncinate fasciculus |  |  |
| AnsaLenticularis | Ansa lenticularis |  |  |
| AnteriorCom | Anterior commissure |  |  |
| LenticularFasc | Lenticular fasciclus |  |  |
| OlfactoryRadiation | olfactory radiation |  |  |
| Mammillary | mammillary body |  |  |
| OpticTract | optic tract |  |  |
| SFG | superior frontal gyrus |  |  |
| MFG | middle frontal gyrus |  |  |
| IFG | inferior frontal gyrus pars triangularis |  |  |
| LFOG | lateral fronto-orbital gyrus |  |  |
| MFOG | middle fronto-orbital gyrus |  |  |
| RG | gyrus rectus |  |  |
| PoCG | postcentral gyrus |  |  |
| PrCG | precentral gyrus |  |  |
| SPG | superior parietal gyrus |  |  |
| SMG | supramarginal gyrus |  |  |
| AG | angular gyrus |  |  |
| PrCu | pre-cuneus |  |  |
| STG | pole of superior temporal gyrus |  |  |
| MTG | pole of middle temporal gyrus |  |  |
| ITG | inferior temporal gyrus |  |  |
| PHG | parahippocampal gyrus |  |  |
| ENT | entorhinal area |  |  |
| FuG | fusiform gyrus |  |  |
| SOG | superior occipital gyrus |  |  |
| MOG | MIDDLE OCCIPITAL GYRUS |  |  |
| IOG | inferior occipital gyrus right |  |  |
| Cu | cuneus |  |  |
| LG | lingual gyrus |  |  |
| rostral | rostral anterior cingulate gyrus |  |  |
| subcallosal | subcallosal anterior cingulate gyrus |  |  |
| subgenual | subgenual anterior cingulate gyrus |  |  |
| dorsal | dorsal anterior cingulate gyrus |  |  |
| PCC | posterior cingulate gyrus |  |  |
| PSTG | posterior superior temporal gyrus |  |  |
| PSMG | posterior middle temporal gyrus |  |  |
| PSIG | posterior inferior temporal gyrus |  |  |

**Radiomics features extraction**

We used the python package PyRadiomics (Version 3.0.1; {<https://pypi.org/project/pyradiomics>}). For the patient level radiomic feature $H$, we define a bag $H=\{\overset{⃑}{h_{1}},\cdots\overset{⃑}{h_{i}},\cdots\overset{⃑}{h_{K}}\}$, where K is the number of lesions in a patient (K is an integer and K$\leq$5) and $\overset{⃑}{h_{i}}$ denotes the radiomic feature vector of the i-th lesion called instances. $\overset{⃑}{h_{i}}=\{f_{i,1},{\ldots,f}_{i,j,}\ldots,f_{i,1454}\}$. The patient-level radiomic feature $H$ is calculated by weighted summation of radiomic features of all delineated lesions as depicted in Equation 1.

The adaptive weight $a_{i}=\left\{ a_{i,1}, a_{i,2},\ldots,a_{i,j},\ldots a_{i,1454} \right\}$ was constructed using a deep learning multi-sample attention model algorithm (Equation 2), where the $w$ and $V$ are network parameters with dimensions of $K\times1454$. The sum of $a_{i}$ for each radiomic feature in an individual patient is 1. After adaptive weighting, the radiomic features (***H***) of a single case was obtained, with the dimension of $1454\times1$.The training process was conducted in the training cohort with a five-fold cross validation strategy. Specifically, the dataset was divided into five folds, where four-folds were used for training and one-fold for validation. The process was repeated five times to ensure every single fold had been used as a validation set once. In each iteration, the radiomic features were extracted from multiple lesions and all 1454 radiomic features were combined as a vector for each lesion. Then, the attention algorithm was applied as described in the previous two steps to generate a patient-level feature set ***H***. It was then fed to a multiple layer perception classifier to generate a fully connected layer with the anticipated bag-level label, sICH was the binary clinical outcome in our study (good prognosis and bad prognosis) and the errors (i.e. function loss) were used to adjust the values of the parameters until a minimized error was reached. The preliminary $w$ and $V$ were determined in the training process and were tuned in the validation fold afterwards.

After five iterations, the mean $w$ and $V$ were used as the final parameters and were kept constant for the rest of the study. Consequently, the adaptive weight coefficients for all lesions and all cases were computed using Equation 2. The patient-level radiomic features were constructed (Supplementary Figure 2) .

Finally, the Lasso feature selection method was applied to select the features with non-zero coefficients from all the weighted features, and 47 features were retained in the end.

$H=\sum_{i=1}^{n=K} a_{i}\overset{⃑}{{\cdot h}_{i}}=\left[ \begin{matrix} \sum_{i=1}^{n=K} a_{i,1}h_{i,1}, & \ldots, & \begin{matrix} \sum_{i=1}^{n=K} a_{i,j}h_{i,j}, & \ldots, & \sum_{i=1}^{n=K} a_{i,1454}h_{i,1454} \end{matrix} \end{matrix} \right]$(1)

$a_{i}=\frac{\exp\left\{ w_{i}^{T}\tanh\left( V_{i}{\overset{⃑}{h_{i}}}^{T} \right) \right\}}{\sum_{l=1}^{K} exp\{w_{i}^{T}tanh(V_{i}{\overset{⃑}{h_{i}}}^{T})\}}$ , where $w_{i}\epsilon R^{1\times1454}$ and $V_{i}\epsilon R^{1\times1454}$ (2)

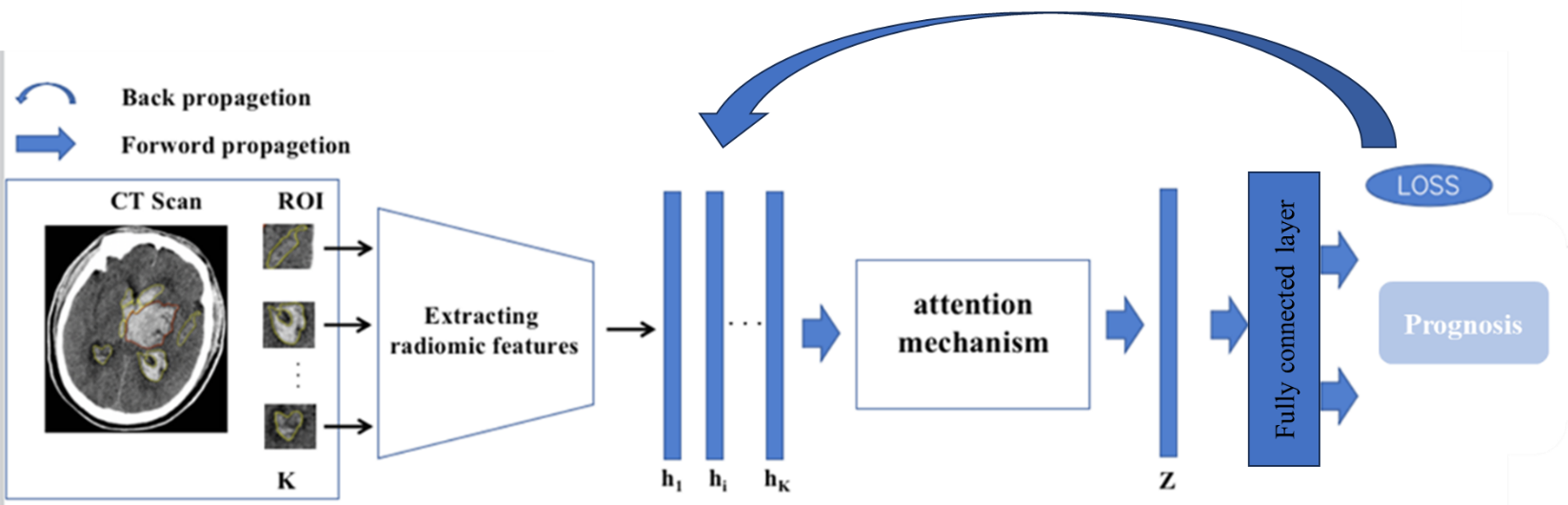
 **Supplementary Figure 2.** Attention mechanism-weighted radiomics feature map

**Supplementary Results**

### **Clinical characteristics**

### Clinical characteristics of development cohort (dataset 1) and external-validation cohorts (dataset 2 and 3) are presented in Supplemental Table1 and 2. There were statistically significant differences in gender and etiology among the three datasets (all p<0.05), statistically significant differences in gender and prognosis between datasets 1 and 2, but no statistically significant differences in gender and prognosis in dataset 3 (*p*>0.05). Stroke history and etiology were statistically significant in the all datasets (all *p*<0.05), stroke history and prognosis were statistically significant in data sets 1 and 3 (all *p*<0.05). Smoking and etiology were statistically significant in dataset 1 (*p*<0.05), but were not statistically significant in etiology and prognosis in the other two datasets (all *p* >0.05). Diabetes mellitus and etiology and prognosis were statistically significant in data sets 1 and 3 (all *p*<0.05), and statistically significant in dataset 2 (all *p*<0.05), but not statistically significant in prognosis (all *p*>0.05). The time from onset to baseline CT and etiology were statistically significant in the three datasets (all *p*<0.05), but there was no statistical significance between the data sets and the prognosis (all *p*<0.05). There were no significant differences in the etiology and prognosis of alcohol consumption and hyperlipidemia among the three datasets (all *p*>0.05) (Supplementary Table 1, 2).

**Supplementary Table2.** **Demographic, Clinical, and Baseline Variables of the Study Population for etiological prediction**

| **Variables** | **Dateset1** | | | |  | **Dateset2** | | | |  | **Dateset3** | | | |
| --- | --- | --- | --- | --- | --- | --- | --- | --- | --- | --- | --- | --- | --- | --- |
|  | **Hypertension** | **Aneurysm** | **Vascular malformation** | ***p* value** |  | **Hypertension** | **Aneurysm** | **Vascular malformation** | ***p* value** |  | **Hypertension** | **Aneurysm** | **Vascular malformation** | ***p* value** |
|  | **(n=337)** | **(n=276)** | **(n=58)** |  |  | **(n= 138)** | **(n= 82)** | **（n=30）** |  |  | **(n= 88)** | **(n= 77)** | **（n=23）** |  |
| Age, median (IQR) | 57  (49-66) | 54  (47-63) | 46  (30.75-57.25) | <0.001* |  | 62  (55-71) | 62  (55-69) | 48  (37.5-54.75) | <0.001* |  | 64.5  (54-74) | 61  (51-68) | 55  (46-59) | 0.001* |
| GCS, median (IQR) | 12  (10-13) | 14  (10-14) | 12  (8.5-14) | <0.001* |  | 11  (8-12) | 13  (10-15) | 12  (10-14) | <0.001* |  | 12  (10-13) | 13  (12-14) | 13  (11-13) | <0.001* |
| Sex, n |  |  |  |  |  |  |  |  |  |  |  |  |  |  |
| Male | 225 (66.8%) | 103 (37.3%) | 34 (58.6%) | <0.001* |  | 98 (71.5%) | 25 (30.5%) | 16 (53.3%) | <0.001* |  | 70 (79.5%) | 33 (42.9%) | 14 (60.9%) | <0.001* |
| Female | 112 (33.2%) | 173 (62.7%) | 24 (41.4%) |  |  | 40 (29.0%) | 57 (69.5%) | 14 (46.7%) |  |  | 18 (20.5%) | 44 (57.1%) | 9 (39.1%) |  |
| Hypertension, n | 337 (100.0%) | 108 (39.1%) | 12 (20.7%) | <0.001* |  | 137 (100.0%) | 53 (64.6%) | 9 (30.0%) | <0.001* |  | 88 (100.0%) | 43 (55.8%) | 7 (30.4%) | <0.001* |
| Diabetes mellitus, n | 48 (14.2%) | 12 (4.3%) | 3 (5.2%) | <0.001* |  | 16 (11.6%) | 2 (2.4%) | 1 (3.3%) | 0.030* |  | 18 (20.5%) | 5 (6.5%) | 1 (4.3%) | 0.012* |
| Hyperlipidemia, n | 17 (5.0%) | 1 (0.4%) | 0 (0.0%) | 0.001* |  | 3 (2.2%) | 1 (1.2%) | 0 (0.0%) | 0.653 |  | 3 (3.4%) | 1 (1.3%) | 1 (4.3%) | 0.608 |
| Smoking, n | 72 (21.4%) | 33 (12.0%) | 6 (10.3%) | 0.003* |  | 32 (23.4%) | 15 (18.3%) | 7 (23.3%) | 0.674 |  | 23 (26.1%) | 14 (18.2%) | 7 (30.4%) | 0.338 |
| Drinking, n | 67 (19.9%) | 48 (17.4%) | 16 (27.6%) | 0.199 |  | 26 (19.0%) | 11 (13.6%) | 5 (16.7%) | 0.525 |  | 19 (21.6%) | 13 (16.9%) | 4 (17.4%) | 0.726 |
| Onset-to-CT time, n |  |  |  |  |  |  |  |  |  |  |  |  |  |  |
| Stage1  (<24h) | 79 (23.4%) | 88 (31.9%) | 20 (34.5%) | 0.014* |  | 114(82.6%) | 73 (89.0%) | 26(86.7%) | <0.001* |  | 78 (88.6%) | 66 (85.7%) | 20 (87%) | <0.001* |
| Stage2  (24h-72h) | 186 (55.2%) | 146 (52.9%) | 26 (44.8%) |  |  | 16(11.6%) | 9(11.0%) | 2(6.7%) |  |  | 10 (11.4%) | 11 (14.3%) | 1 (4.3%) |  |
| Stage3  (3-7d) | 57 (16.9%) | 41 (14.9%) | 10 (17.2%) |  |  | 6 (4.3%) | 0(0.0%) | 2(6.7%) |  |  | 0 (0.0%) | 0 (0.0%) | 1 (4.3%) |  |
| Stage  (>7d) | 15 (4.5%) | 1 (0.4%) | 2 (3.4%) |  |  | 2(1.4%) | 0 (0.0%) | 0(0.0%) |  |  | 0 (0.0%) | 0 (0.0%) | 1 (4.3%) |  |
| History of stroke, n | 29 (8.6%) | 2 (0.7%) | 2 (3.4%) | <0.001* |  | 31 (22.5%) | 6 (7.3%) | 3 (10.0%) | 0.008* |  | 11 (12.5%) | 1 (1.3%) | 7 (30.4%) | <0.001* |

IQR= interquartile range; GCS= Glasgow Coma Scale; *Significant difference (P<0.05)

**Supplementary Table3. Demographic, Clinical, and Baseline Variables of the Study Population for prognosis prediction**

| **Variables** | **Dateset1** | | |  | **Dateset2** | | |  | **Dateset3** | | |
| --- | --- | --- | --- | --- | --- | --- | --- | --- | --- | --- | --- |
|  | **mRS<3** | **mRS>=3** | ***p* value** |  | **mRS<3** | **mRS>=3** | ***p value*** |  | **mRS<3** | **mRS>=3** | ***p value*** |
|  | **(n=448)** | **(n=250)** |  |  | **(n= 163)** | **（n=106）** |  |  | **(n= 110)** | **（n=85）** |  |
| Age, median (IQR) | 52 | 61.5 | <0.001* |  | 59 | 59 | <0.001* |  | 55.5 | 68 | <0.001* |
|  | (46-61.25) | (52 -69) |  |  | (53-70) | (53-70) |  |  | (50-64.75) | (58-75) |  |
| GCS, median (IQR) | 14 | 10 | <0.001* |  | 13 | 10 | 0.008 |  | 13 | 11 | <0.001* |
|  | (11.75-14) | (8-12) |  |  | (11-14) | (7.25-12) |  |  | (13-14) | (9-12) |  |
| Sex, n |  | | |  |  |  |  |  |  |  |  |
| Male | 218(48.7%) | 101(40.4%) | 0.036 |  | 85 (52.1%) | 69 (65.1%) | 0.036 |  | 65 (59.1%) | 57 (67.1%) | 0.254 |
| Female | 230(51.3%) | 149(59.6%) |  |  | 78 (47.9%) | 37 (34.9%) |  |  | 45 (40.9%) | 28 (32.9%) |  |
| Hypertension, n | 243(54.2%) | 185(74.0%) | <0.001* |  | 109(66.9%) | 85 (80.2%) | 0.015 |  | 64 (58.2%) | 71 (83.5%) | <0.001* |
| Diabetes mellitus, n | 29 (6.5%) | 36 (14.4%) | <0.001* |  | 13 (8.0%) | 7 (6.6%) | 0.675 |  | 8 (7.3%) | 17 (20.0%) | 0.008 |
| Hyperlipidemia, n | 9 (2.0%) | 9 (3.6%) | 0.204 |  | 3 (1.8%) | 1 (0.9%) | 0.937 |  | 3 (2.7%) | 3 (2.7%) | 1 |
| Smoking, n | 68 (15.2%) | 46 (18.4%) | 0.27 |  | 30 (18.4%) | 30 (28.3%) | 0.057 |  | 27 (24.5%) | 19 (22.4%) | 0.721 |
| Drinking, n | 78 (17.4%) | 56 (22.4%) | 0.109 |  | 23 (14.2%) | 24 (22.6%) | 0.076 |  | 22 (20.0%) | 16 (18.8%) | 0.837 |
| Pathogenesis, n |  |  |  |  |  |  |  |  |  |  |  |
| Hypertension | 166(37.1%) | 171(68.4%) | <0.001* |  | 71 (43.6%) | 66 (62.3%) | 0.007 |  | 35 (31.8%) | 53 (62.4%) | <0.001* |
| Arterial aneurysm | 219(48.9%) | 57 (22.8%) |  |  | 59 (36.2%) | 23 (21.7%) |  |  | 55 (50.0%) | 22 (25.9%) |  |
| Vascular malformation | 36 (8.0%) | 22 (8.8%) |  |  | 17 (10.4%) | 13 (12.3%) |  |  | 17 (15.5%) | 6 (7.1%) |  |
| Unknown | 27 (6.0%) | 0 (0.0%) |  |  | 16 (9.8%) | 4 (3.8%) |  |  | 3 (2.7%) | 4 (4.7%) |  |
| Onset-to-CT time, n |  |  |  |  |  |  |  |  |  |  |  |
| Stage1 | 116 | 79 | 0.263 |  | 126 | 92 | 0.155 |  | 92(83.6%) | 78 (91.8%) | 0.262 |
| (<24h) |  |  |  |  |  |  |  |  |  |  |  |
| Stage2 | 242 | 371 |  |  | 22 | 12 |  |  | 16 (14.5%) | 7 (8.2%) |  |
| (24h-72h) |  |  |  |  |  |  |  |  |  |  |  |
| Stage3 | 79 | 113 |  |  | 12 | 2 |  |  | 1 (0.9%) | 0 (0.0%) |  |
| (3-7d) |  |  |  |  |  |  |  |  |  |  |  |
| Stage | 11 | 19 |  |  | 3 | 0 |  |  | 1 (0.9%) | 0 (0.0%) |  |
| (>7d) |  |  |  |  |  |  |  |  |  |  |  |
| History of stroke, n | 13 (2.9%) | 21 (8.4%) | <0.001* |  | 19 (11.7%) | 21 (19.8%) | 0.066 |  | 7 (6.4%) | 12 (14.1%) | 0.07 |

IQR= interquartile range; GCS= Glasgow Coma Scale; mRS= the modified Rankin Scale; *Significant difference (p<0.05)

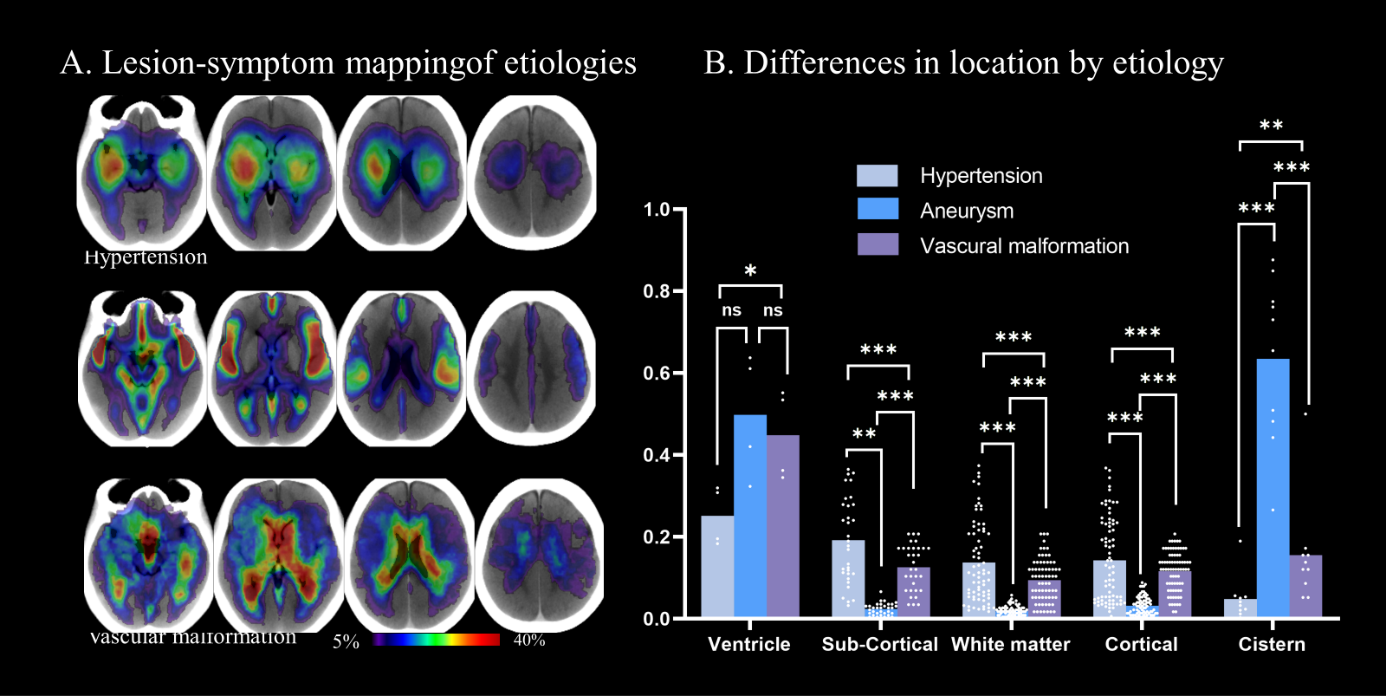

**Supplementary Figure 3. Association between hematoma location and etiology in the dateset1 (n = 671).** (A) Probability maps depicting the distribution of hematoma location in patients. The location patterns associated with hypertension, aneurysm, and vascular malformation are illustrated, with the frequency of hemorrhage location graded according to color. The gradient scale ranges from 5% to 40%. (B) Paired sample ANOVA was used to compare the involvement rate of each region in each intracranial structure. *: p < 0.05, **: p < 0.01, ***: p < 0.001, ns: p > 0.05.

**Supplementary**
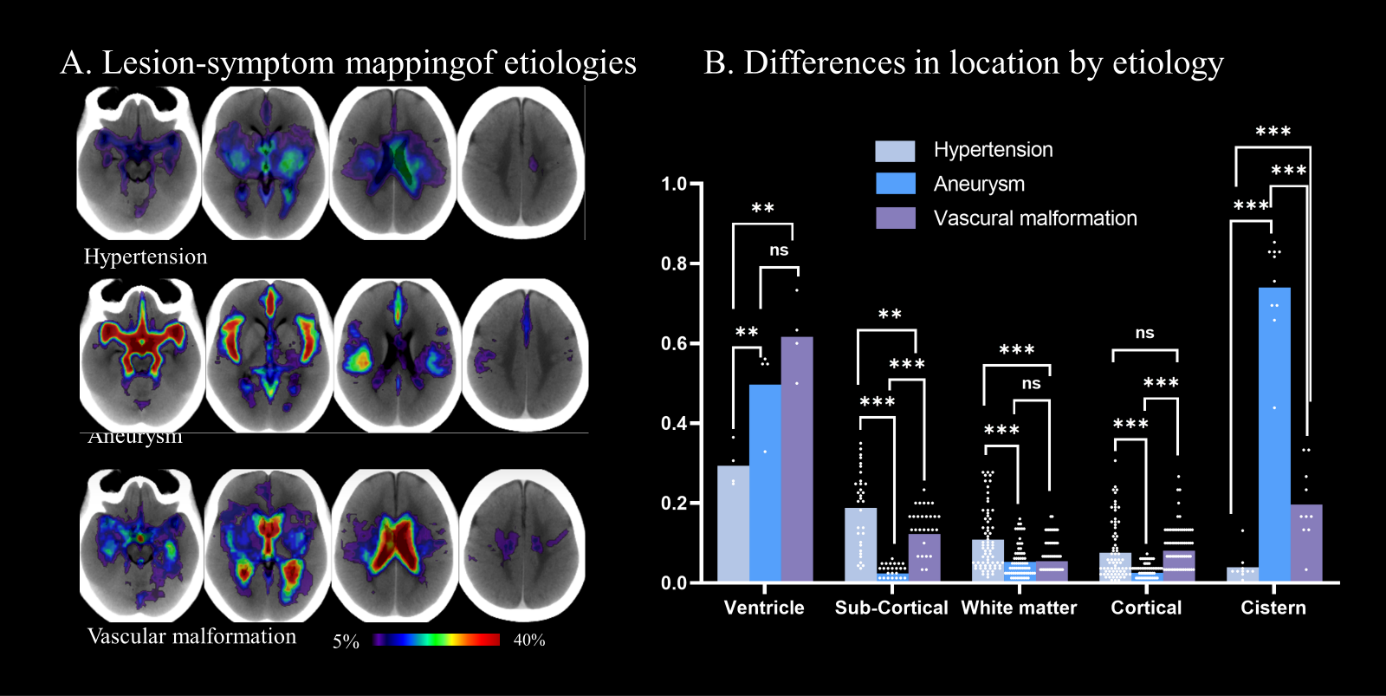
**Figure 4. Association between hemorrhage location and etiology in the dateset2 (n = 250).** (A) Probability maps depicting the distribution of hematoma location in patients. The location patterns associated with hypertension, aneurysm, and vascular malformation are illustrated, with the frequency of hemorrhage location graded according to color. The gradient scale ranges from 5% to 40%. (B) Paired sample ANOVA was used to compare the involvement rate of each region in each intracranial structure. *: p < 0.05, **: p < 0.01, ***: p < 0.001, ns: p > 0.05.

L

**Supplementary**
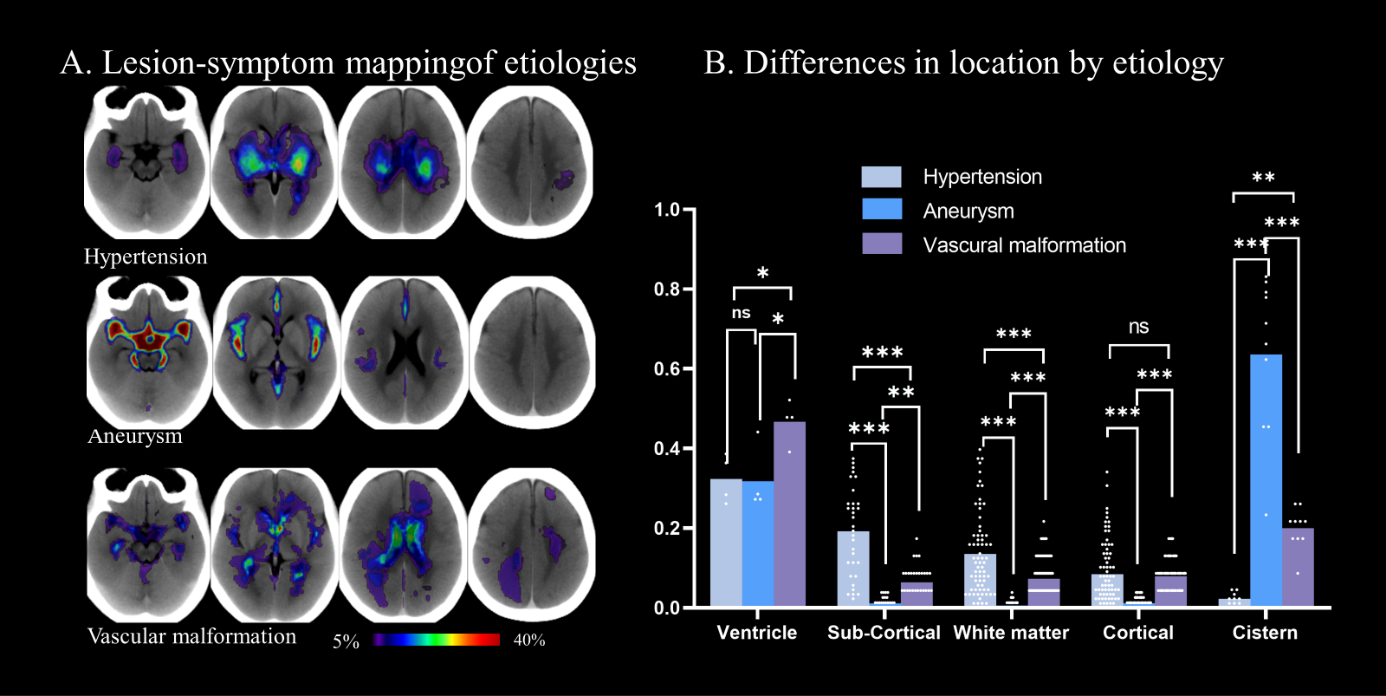
**Figure 5. Lesion-symptom mapping of hemorrhage location in the dateset3 (n = 188).** (A) Probability maps depicting the distribution of hematoma location in patients. The location patterns associated with hypertension, aneurysm, and vascular malformation are illustrated, with the frequency of hemorrhage location graded according to color. The gradient scale ranges from 5% to 40%. (B) Paired sample ANOVA was used to compare the involvement rate of each region in each intracranial structure. *: p < 0.05, **: p < 0.01, ***: p < 0.001, ns: p > 0.05.

**Supplementary**
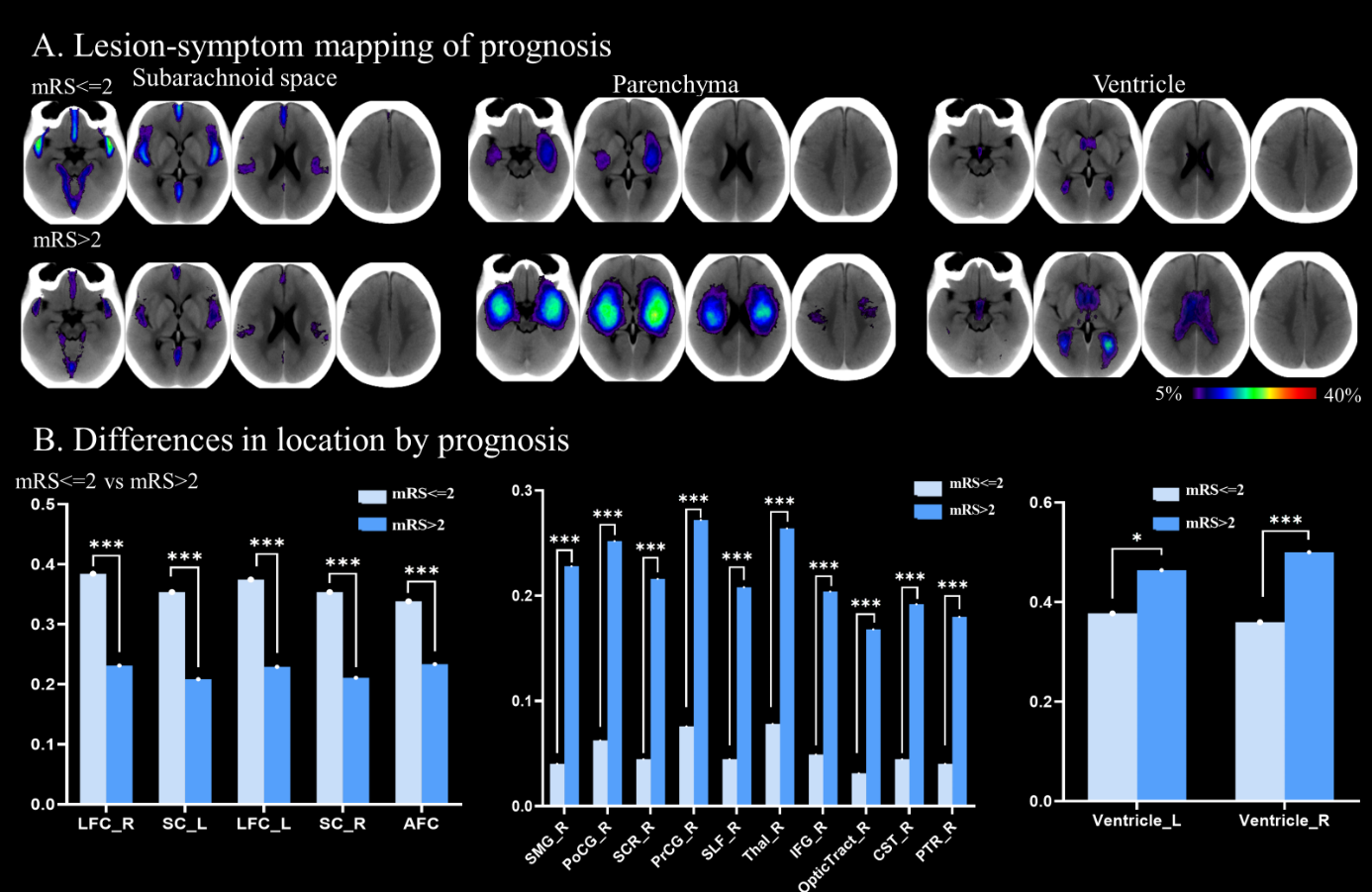
**Figure 6.** **Association between hemorrhage location and prognosis in dataset1 (n = 698). (A)** Probability maps depicting the distribution of hematoma location in different prognosis. The location patterns associated with subarachnoid space, parenchyma and ventricle are illustrated, with the frequency of hematoma location graded according to color. The gradient scale ranges from 5% to 40%. (B) Frequency plots of the top five cisterns, ten parenchyma and ventricles with group difference, ranked according to chi-square values. *: p < 0.05, **: p < 0.01, ***: p < 0.001. Abbreviations of intracranial region see Supplementary Table 1.

L

**Supplementary**
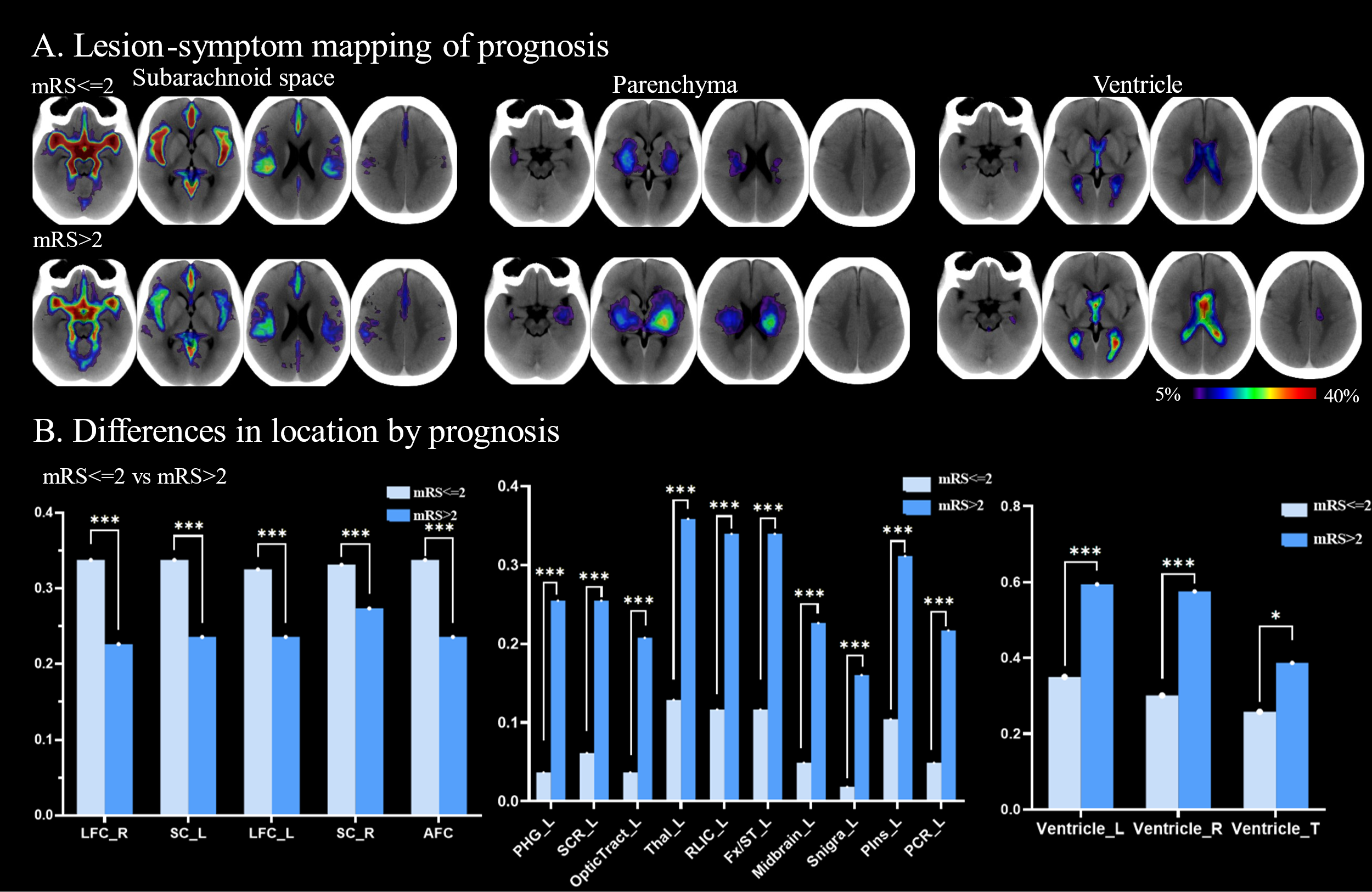
**Figure 7. Association between hemorrhage location and prognosis in dataset2 (n = 269).** (A) Probability maps depicting the distribution of hematoma location in different prognosis. The location patterns associated with subarachnoid space, parenchyma and ventricle are illustrated, with the frequency of hematoma location graded according to color. The gradient scale ranges from 5% to 40%. (B) Frequency plots of the top five cisterns, ten parenchyma and ventricles with group difference, ranked according to chi-square values. *: p < 0.05, **: p < 0.01, ***: p < 0.001. Abbreviations of intracranial region see Supplementary Table 1.

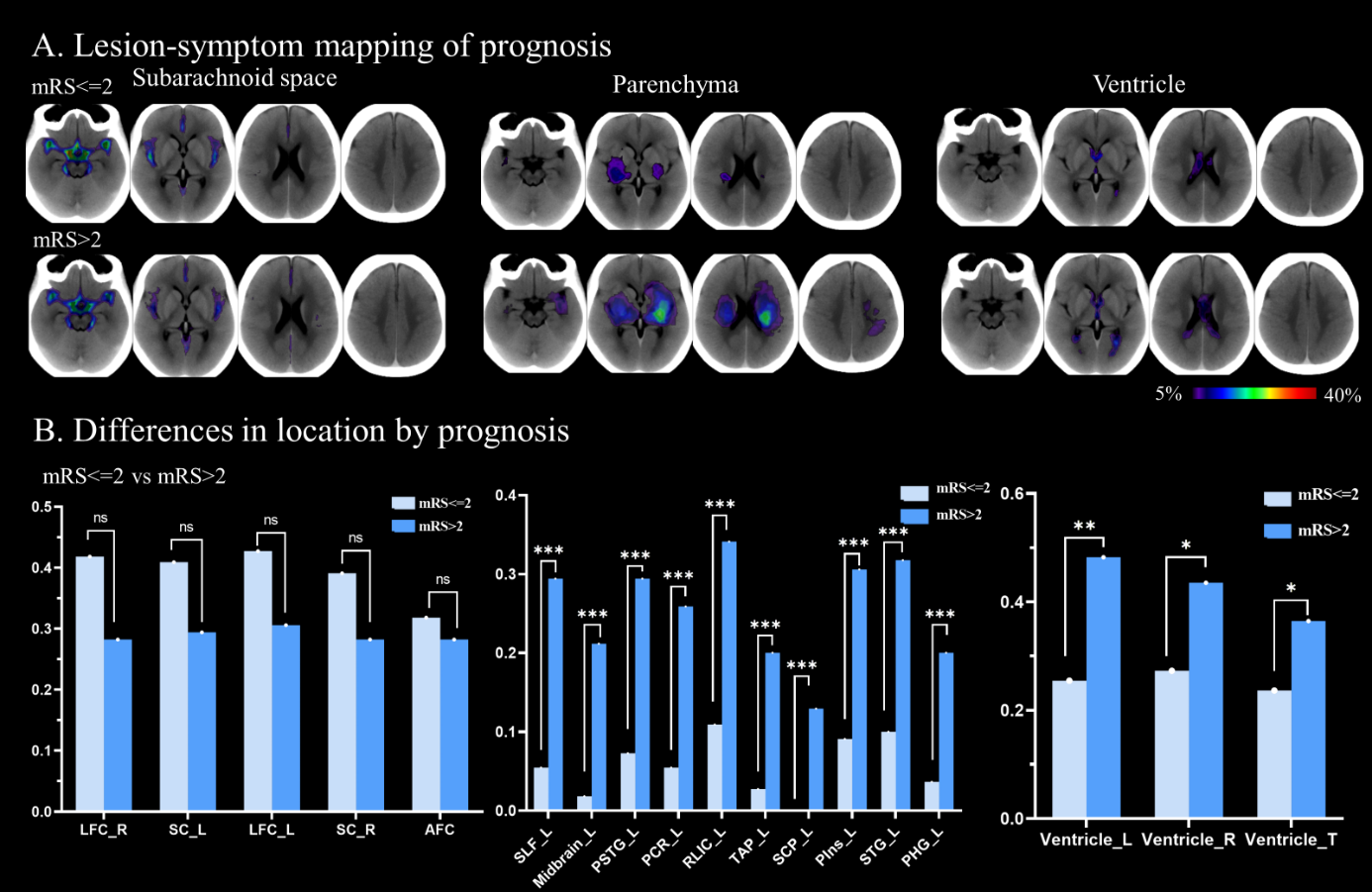

**Supplementary Figure 8. Association between hemorrhage location and prognosis in dataset3 (n = 195).** **(A)** Probability maps depicting the distribution of hematoma location in different prognosis. The location patterns associated with subarachnoid space, parenchyma and ventricle are illustrated, with the frequency of hematoma location graded according to color. The gradient scale ranges from 5% to 40%. **(B)** Frequency plots of the top five cisterns, ten parenchyma and ventricles with group difference, ranked according to chi-square values. *: p < 0.05, **: p < 0.01, ***: p < 0.001, ns: p > 0.05. Abbreviations of intracranial region see Supplementary Table 1.

L

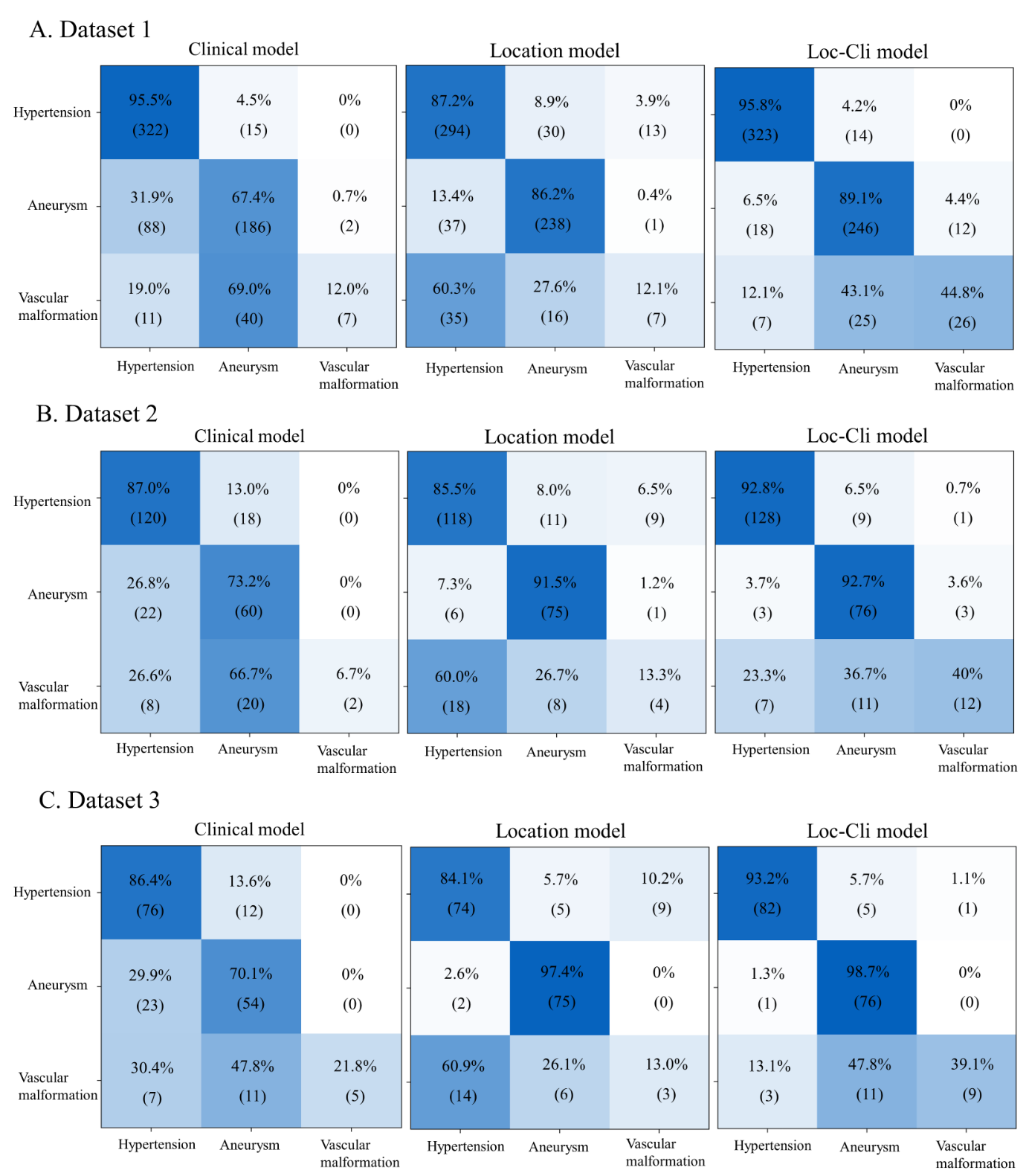
**Supplementary Figure 9. The confusion matrix of etiology models**. (A-C) The confusion matrix demonstrates the classification capability of the clinical, location and loc-cli models for predicting etiology in dataset 1,2,3. where columns are predicted classes and rows are ground truth.

Loc-Cli= location-clinical

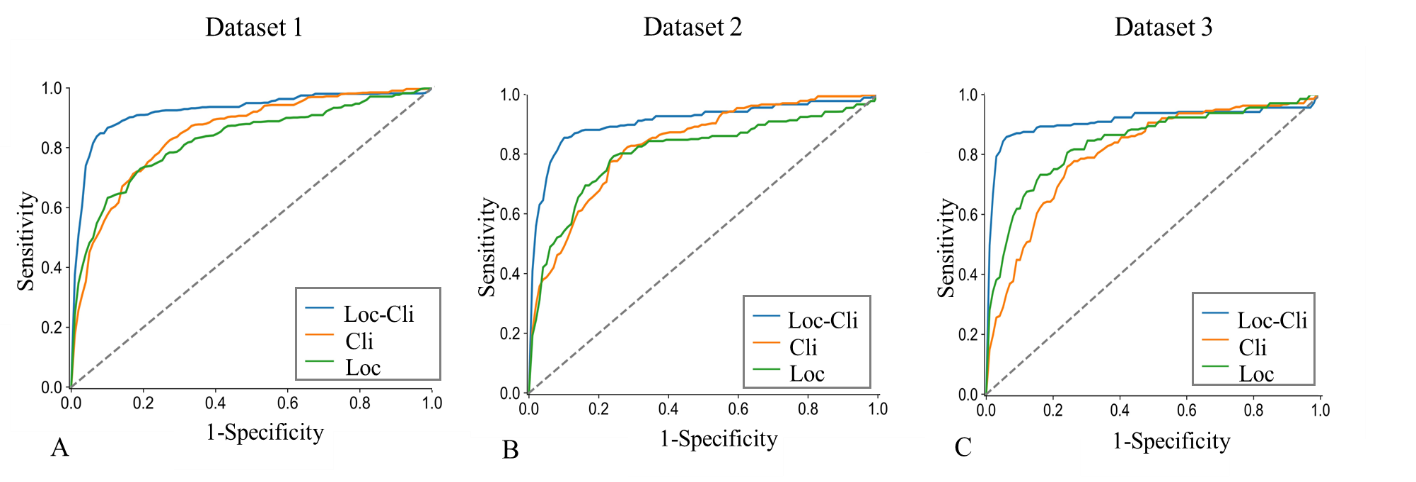
 **Supplementary Figure 10. Etiology of the ROC curves.** ROC curves of etiology models of each dataset.

ROC= Receiver Operating Characteristic

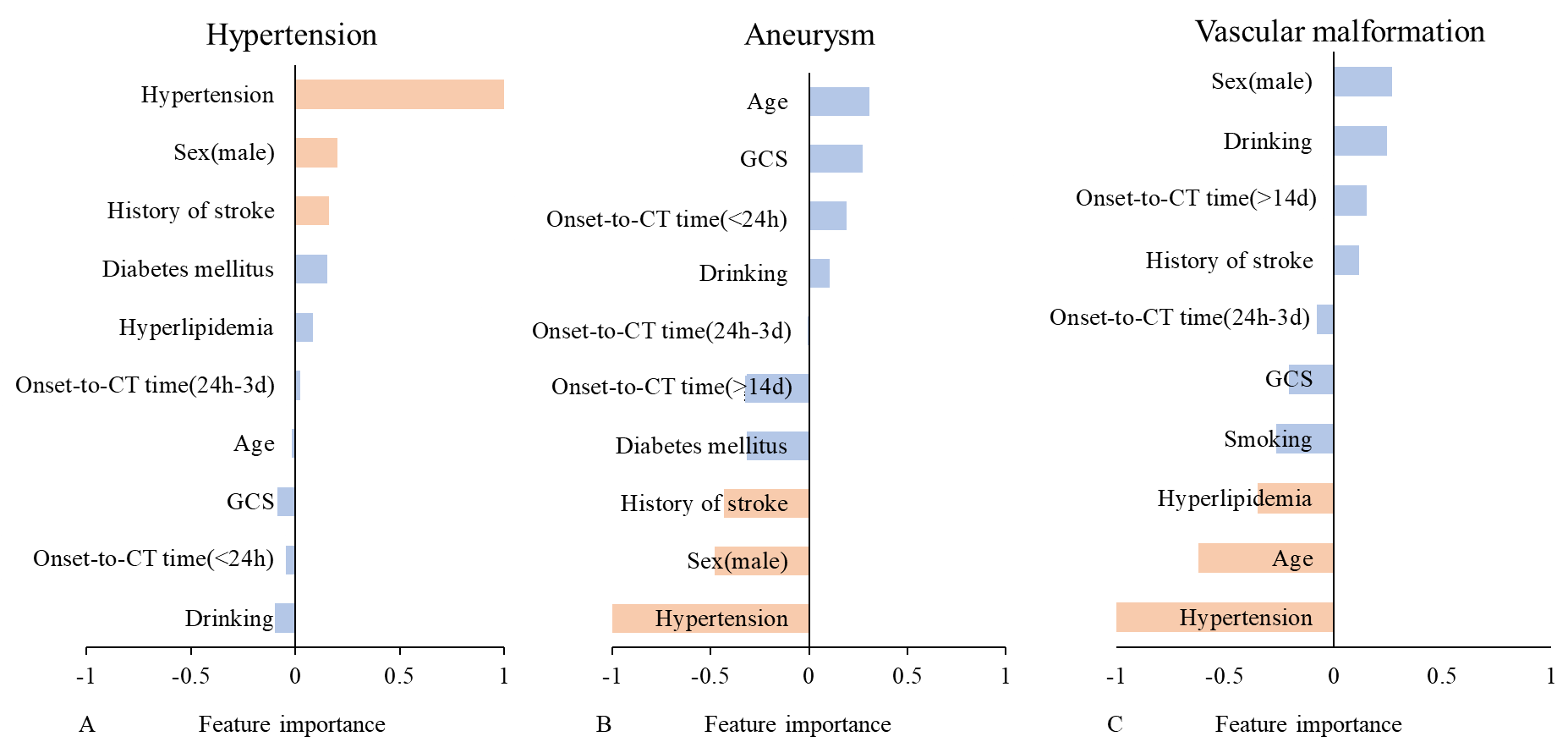
**Supplementary Figure 11**. **Feature importance of etiology clinical models.** (A-C) Show the top ten clinical features most associated with hypertension, aneurysm, vascular malformation, red represents the three features that contribute the most. Positive values are positively contribution and negative values are negatively contribution. GCS= Glasgow Coma Scale;

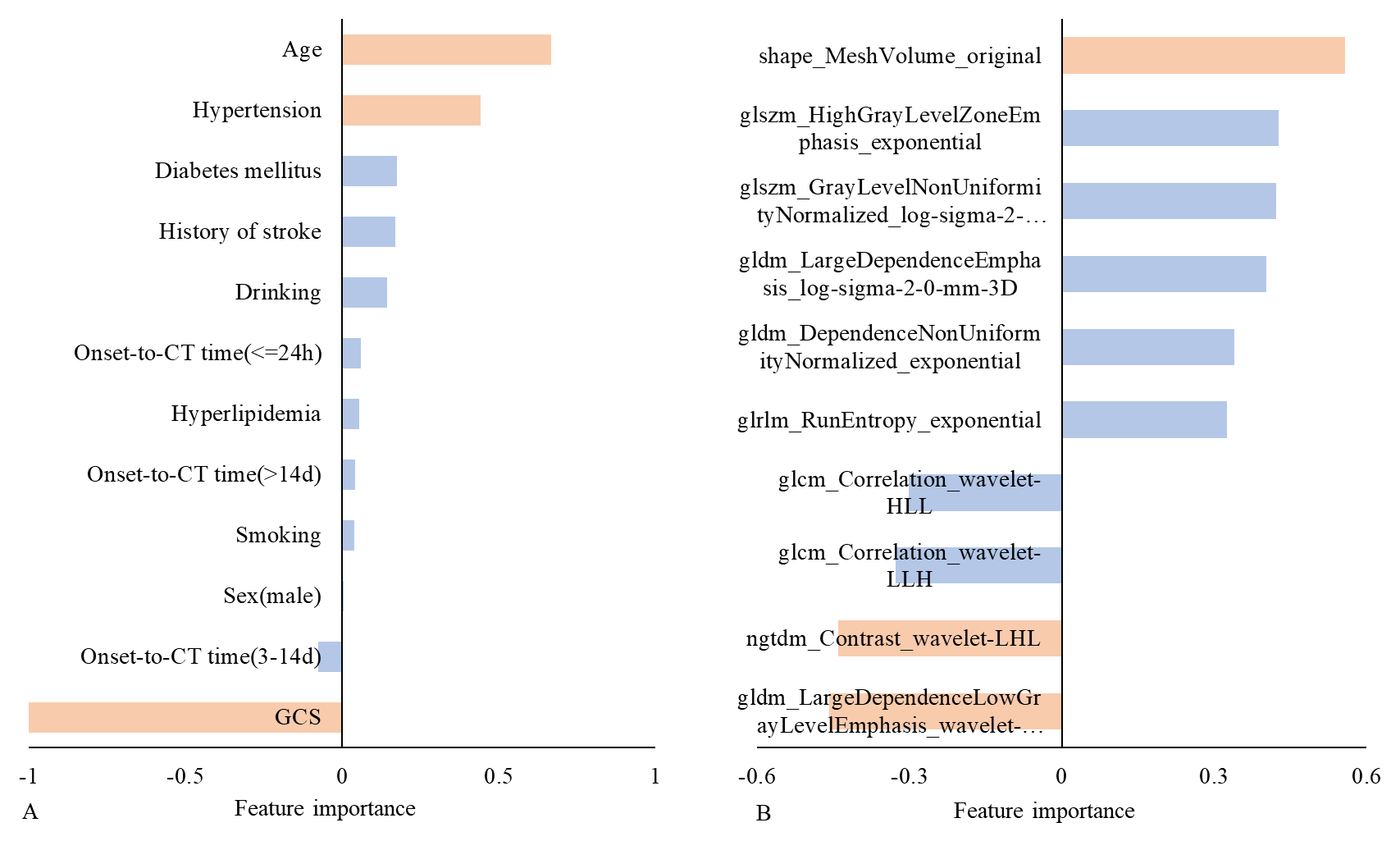
**Supplementary Figure 12. Feature importance of prognosis models.** (A) The top ten clinical features most associated with prognosis. (B) The top ten radiomics features most associated with prognosis, red represents the three features that contribute the most. Positive values are positively contribution and negative values are negatively contribution.

GCS= Glasgow Coma Scale;

**Supplementary Table 4.** **Performance of clinical model, location model, radiomics model and fusion model prediction task model of all datasets**

|  | **Dateset1** | | | |  | **Dateset2** | | | |  | **Dateset3** | | | |
| --- | --- | --- | --- | --- | --- | --- | --- | --- | --- | --- | --- | --- | --- | --- |
|  | AUC | Accuracies | Specificity | Sensitivity |  | AUC | Accuracies | Specificity | Sensitivity |  | AUC | Accuracies | Specificity | Sensitivity |
|  | (95% CI) |  |  |  |  | (95% CI) |  |  |  |  | (95% CI) |  |  |  |
| **Etiology** |  |  |  |  |  |  |  |  |  |  |  |  |  |  |
| Clinical model | 0.848 | 0.767 | 0.858 | 0.583 |  | 0.831 | 0.728 | 0.835 | 0.556 |  | 0.806 | 0.718 | 0.831 | 0.594 |
|  | (0.828-0.867) |  |  |  |  | (0.797-0.865) |  |  |  |  | (0.765-0.847) |  |  |  |
| Location model | 0.826 | 0.803 | 0.882 | 0.658 |  | 0.807 | 0.788 | 0.876 | 0.634 |  | 0.842 | 0.809 | 0.895 | 0.649 |
|  | (0.805-0.847) |  |  |  |  | (0.771-0.842) |  |  |  |  | (0.804-0.879) |  |  |  |
| Loc-Cli model | 0.923 | 0.887 | 0.936 | 0.766 |  | 0.909 | 0.864 | 0.924 | 0.752 |  | 0.912 | 0.893 | 0.939 | 0.774 |
|  | (0.909-0.938) |  |  |  |  | (0.883-0.935) |  |  |  |  | (0.883-0.941) |  |  |  |
| **Prognosis** |  |  |  |  |  |  |  |  |  |  |  |  |  |  |
| Clinical model | 0.799 | 0.716 | 0.670 | 0.800 |  | 0.772 | 0.647 | 0.528 | 0.830 |  | 0.762 | 0.758 | 0.848 | 0.656 |
|  | (0.766-0.832) |  |  |  |  | (0.717 -0.821) |  |  |  |  | (0.666-0.795) |  |  |  |
| Location model | 0.785 | 0.705 | 0.652 | 0.800 |  | 0.725 | 0.647 | 0.521 | 0.840 |  | 0.777 | 0.773 | 0.838 | 0.699 |
|  | (0.753 -0.815) |  |  |  |  | (0.668 -0.778) |  |  |  |  | (0.708-0.830) |  |  |  |
| Radiomics model | 0.839 | 0.769 | 0.777 | 0.756 |  | 0.831 | 0.751 | 0.755 | 0.745 |  | 0.840 | 0.783 | 0.829 | 0.731 |
|  | (0.810 -0.866) |  |  |  |  | (0.780 -0.873) |  |  |  |  | (0.782-0.889) |  |  |  |
| Loc-Cli-Rad model | 0.895 | 0.855 | 0.888 | 0.796 |  | 0.874 | 0.829 | 0.865 | 0.774 |  | 0.850 | 0.879 | 0.905 | 0.849 |
|  | (0.869 -0.916) |  |  |  |  | (0.828 -0.911) |  |  |  |  | (0.790-0.895) |  |  |  |

AUC= Area under the curve; Loc-Cli model = Location- Clinical model; Loc-Cli-Rad model= Location- Clinical- Radiomics model

**Supplementary Table 5. Delong analysis of AUC values of models**

| **Delong test-etiology** | | | | | | | | | | | |
| --- | --- | --- | --- | --- | --- | --- | --- | --- | --- | --- | --- |
|  | **Dateset1** | | |  | **Dateset2** | | |  | **Dateset3** | | |
|  | AUC  (Cli) | AUC  (Loc-Cli) | p value |  | AUC  (Cli) | AUC  (Loc-Cli) | p value |  | AUC  (Cli) | AUC  (Loc-Cli) | p value |
| **Etiology** | 0.848 | 0.923 | < 0.0001 |  | 0.831 | 0.909 | 0.0001 |  | 0.806 | 0.912 | 0.0002 |
| **Delong test-prognosis** | | | | | | | | | | | |
|  | AUC  (Cli) | AUC  (Loc-Cli-Rad) | p value |  | AUC  (Cli) | AUC  (Loc-Cli-Rad) | p value |  | AUC  (Cli) | AUC  (Loc-Cli-Rad) | p value |
| **Prognosis** | 0.799 | 0.895 | < 0.0001 |  | 0.772 | 0.874 | 0.003 |  | 0. 762 | 0.850 | 0.004 |

Cli = clinical; Loc-Cli=location-clinical; Loc-Cli-Rad= location-clinical-radiomics; AUC= AUC= Area under the curve.
